# Genomics and epidemiology of a novel SARS-CoV-2 lineage in Manaus, Brazil

**DOI:** 10.1101/2021.02.26.21252554

**Authors:** Nuno R. Faria, Thomas A. Mellan, Charles Whittaker, Ingra M. Claro, Darlan da S. Candido, Swapnil Mishra, Myuki A. E. Crispim, Flavia C. Sales, Iwona Hawryluk, John T. McCrone, Ruben J. G. Hulswit, Lucas A. M. Franco, Mariana S. Ramundo, Jaqueline G. de Jesus, Pamela S. Andrade, Thais M. Coletti, Giulia M. Ferreira, Camila A. M. Silva, Erika R. Manuli, Rafael H. M. Pereira, Pedro S. Peixoto, Moritz U. Kraemer, Nelson Gaburo, Cecilia da C. Camilo, Henrique Hoeltgebaum, William M. Souza, Esmenia C. Rocha, Leandro M. de Souza, Mariana C. de Pinho, Leonardo J. T Araujo, Frederico S. V. Malta, Aline B. de Lima, Joice do P. Silva, Danielle A. G. Zauli, Alessandro C. de S. Ferreira, Ricardo P Schnekenberg, Daniel J. Laydon, Patrick G. T. Walker, Hannah M. Schlüter, Ana L. P. dos Santos, Maria S. Vidal, Valentina S. Del Caro, Rosinaldo M. F. Filho, Helem M. dos Santos, Renato S. Aguiar, José L. P. Modena, Bruce Nelson, James A. Hay, Melodie Monod, Xenia Miscouridou, Helen Coupland, Raphael Sonabend, Michaela Vollmer, Axel Gandy, Marc A. Suchard, Thomas A. Bowden, Sergei L. K. Pond, Chieh-Hsi Wu, Oliver Ratmann, Neil M. Ferguson, Christopher Dye, Nick J. Loman, Philippe Lemey, Andrew Rambaut, Nelson A. Fraiji, Maria do P. S. S. Carvalho, Oliver G. Pybus, Seth Flaxman, Samir Bhatt, Ester C. Sabino

## Abstract

Cases of SARS-CoV-2 infection in Manaus, Brazil, resurged in late 2020, despite high levels of previous infection there. Through genome sequencing of viruses sampled in Manaus between November 2020 and January 2021, we identified the emergence and circulation of a novel SARS-CoV-2 variant of concern, lineage P.1, that acquired 17 mutations, including a trio in the spike protein (K417T, E484K and N501Y) associated with increased binding to the human ACE2 receptor. Molecular clock analysis shows that P.1 emergence occurred around early November 2020 and was preceded by a period of faster molecular evolution. Using a two-category dynamical model that integrates genomic and mortality data, we estimate that P.1 may be 1.4–2.2 times more transmissible and 25-61% more likely to evade protective immunity elicited by previous infection with non-P.1 lineages. Enhanced global genomic surveillance of variants of concern, which may exhibit increased transmissibility and/or immune evasion, is critical to accelerate pandemic responsiveness.

**One-Sentence Summary:** We report the evolution and emergence of a SARS-CoV-2 lineage of concern associated with rapid transmission in Manaus.

---

Brazil has experienced high mortality during the COVID-19 pandemic, recording >250,000 deaths and >10 million reported cases, as of February 2020. SARS-CoV-2 infection and disease burden have been highly variable across the country, with Amazonas state in north Brazil being the worst-affected region (*1*). Serological surveillance of blood donors in Manaus, the capital city of Amazonas and the largest city in the Amazon region, has suggested >67% cumulative attack rates by October 2020 (*2*). Similar but slightly lower seroprevalences have also been reported for cities in neighbouring regions (*3, 4*). However, the level of previous infection in Manaus was clearly not sufficient to prevent a rapid resurgence in SARS-CoV-2 transmission and mortality there during late 2020 and early 2021 (*5*), which has placed a significant pressure on the city’s healthcare system.

Here, we show that the second wave of infection in Manaus was associated with the emergence and rapid spread of a new SARS-CoV-2 lineage of concern, named lineage P.1. The lineage carries a unique constellation of mutations, including several that have been previously determined to be of virological importance (*6-10*) and which are located in the spike protein receptor binding domain (RBD), the region of the virus involved in recognition of the angiotensin-converting enzyme-2 receptor cell surface receptor (*11*). Using genomic data, structure-based mapping of mutations of interest onto the spike protein, and dynamical epidemiology modelling of genomic and mortality data, we investigate the emergence of the P.1 lineage and explore epidemiological explanations for the resurgence of COVID-19 in Manaus.

## Identification and nomenclature of a novel P.1 lineage in Manaus

In late 2020, two SARS-CoV-2 lineages of concern were discovered through genomic surveillance, both characterised by sets of significant mutations: lineage B.1.351, first reported in South Africa (*12*) and lineage B.1.1.7, detected in the United Kingdom (*13*). Both variants have transmitted rapidly in the countries where they were discovered and spread to other regions (*14, 15*). Analyses indicate B.1.1.7 has higher transmissibility than previously circulating lineages in the UK (*13*).

Following a rapid increase in hospitalizations in Manaus caused by severe acute respiratory infection in Dec 2020 (Fig. 1A), we focused ongoing SARS-CoV-2 genomic surveillance (*2, 16-20*) on recently collected samples from the city (see **Materials and Methods**). Prior to this, only seven SARS-CoV-2 genome sequences from Amazonas state were publicly available (SARS-CoV-2 was first detected in Manaus on 13 March 2020) (*17, 21*). We sequenced SARS-CoV-2 genomes from 184 samples from patients seeking COVID-19 testing in two diagnostic laboratories in Manaus between November and December 2020, using the ARTIC V3 multiplexed amplicon scheme (*22*) and the MinION sequencing platform. As partial genome sequences can provide useful epidemiological information, particularly regarding virus genetic diversity and lineage composition (*23*), we harnessed information from partial (*n*=41, 25-75% genome coverage), as well as near-complete (*n*=95, 75-95%) and complete (*n*=48, ≥95%) sequences from Manaus (fig. S1-S3), together with other available and published genomes from Brazil for context (**Data S3**). Pango lineages were classified using the Pangolin (*24*) software tool (http://pangolin.cog-uk.io/) and standard phylogenetic analysis using complete reference genomes.

**Fig. 1.**
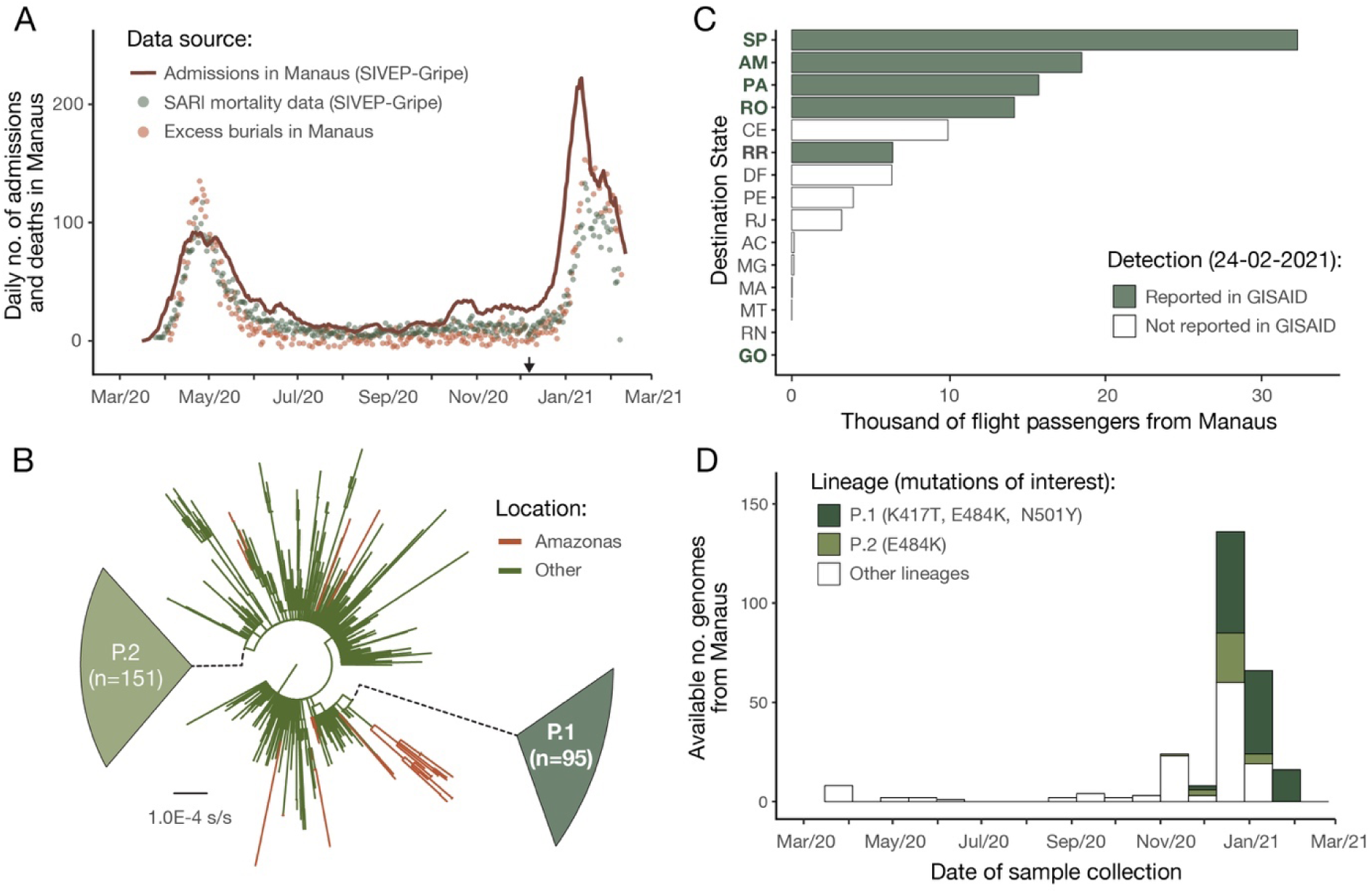
SARS-CoV-2 epidemiological, diagnostic, genomic and mobility data from Manaus. (**A**) Dark solid line shows the 7-day rolling average of the COVID-19 confirmed and suspected daily time series of hospitalisations in Manaus. Admissions in Manaus are from Fundação de Vigilância em Saúde do Amazonas (*75*). Green dots represent daily severe acute respiratory mortality records from the SIVEP-Gripe (*Sistema de Informação de Vigilância Epidemiológica da Gripe*) database (*71*). SARI = severe acute respiratory infections. Excess burial records based on data from Manaus Mayor’s office are shown in red dots for comparison (see Materials and Methods). The arrow denotes 6 December 2020, the date of the first P.1 case identified in Manaus by our study. (**B**) Maximum likelihood tree (*n*=974) with B.1.1.28, P.1 and P.2 sequences, with collapsed views of P.1 and P.2 clusters and highlighting other sequences from Manaus, Brazil). Ancestral branches leading to P.1 and P.2 are shown as dashed lines. See fig. S3 for a more detail phylogeny. Scale bar is shown in units of nucleotide substitutions per site (s/s). (**C**) Number of air travel passengers from Manaus to all states in Brazil was obtained from National Civil Aviation Agency of Brazil (www.gov.br/anac). The ISO 3166-2:BR codes of the states with genomic reports of P.1 (GISAID (*76*), as of 24 Feb 2021), are shown in bold. An updated list of GISAID genomes and reports of P.1 worldwide is available at https://cov-lineages.org/global_report_P.1.html. (**D**) Number of genome sequences from Manaus belonging to lineages of interest (see Materials and Methods); spike mutations of interest are denoted.

Our early data indicated the presence of a novel SARS-CoV-2 lineage in Manaus containing 17 amino acid changes (including 10 in the spike protein), 3 deletions, 4 synonymous mutations and a 4 nucleotide insertion compared to the most closely related available sequence (GISAID ID: EPI_ISL_722052) (*25*) (Fig. 1B). This lineage was given a new designation, P.1, on the basis that (*i*) it is phylogenetically and genetically distinct from ancestral viruses, (*ii*) associated with rapid spread in a new area, and (*iii*) carries a constellation of mutations that may have phenotypic relevance (*24*). Phylogenetic analysis indicated that P.1, and another lineage, P.2 (*17*), were descendants of lineage B.1.1.28 that was first detected in Brazil in early March 2020 (Fig 1B). Our preliminary results were shared with local teams on 10 Jan 2021 and published online on 12 Jan 2021 (*25*). Concurrently, cases of SARS-CoV-2 P.1 infection were reported in Japan in travellers from Amazonas (*26*). As of 24 Feb 2021, P.1 had been confirmed in 6 Brazilian states, which in total received >92,000 air passengers from Manaus in November 2020 (Fig. 1C). Genomic surveillance first detected lineage P.1 on 6 December 2020 (Fig. 1A), after which the frequency of P.1 relative to other lineage increased rapidly in the tested samples from Manaus (Fig. 1D; lineage frequency information can be found in fig. S2).

## Dating the emergence of the P.1 lineage

We next sought to understand the emergence and evolution of lineage P.1 using molecular clock phylogenetics (*23*). We first regressed root-to-tip genetic distances against sequence sampling dates (*27*) for the P.1, P.2, and B.1.1.28 lineages separately. This exploratory analysis revealed similar evolutionary rates within each lineage, but greater root-to-tip distances for P.1 compared to B.1.1.28 (fig. S4), suggesting that the emergence of P.1 was preceded by a period of faster molecular evolution. The B.1.1.7 lineage also exhibits such evolution (*13*), which was hypothesised to have occurred in a chronically infected or immunocompromised patient (*28, 29*).

In order to date the emergence of P.1 while accounting for a faster evolutionary rate along its ancestral branch, we used a local molecular clock model (*30*) with a flexible non-parametric demographic tree prior (*31*). Using this approach, we estimate the date of the common ancestor of the P.1 lineage to around 6 Nov 2020 (95% Bayesian credible interval, BCI, 9 Oct to 30 Nov 2020). This is approximately one month prior to the resurgence in SARS-CoV-2 confirmed cases in Manaus (Fig. 1A, Fig.2). The P.1 sequences formed a single well-supported group (posterior probability=1.00) that clustered most closely with B.1.1.28 sequences from Manaus (“*AM*” in Fig. 2), suggesting P.1 emerged there. Further, the local clock model statistically confirms a higher evolutionary rate for the branch immediately ancestral to lineage P.1 compared to that for lineage B.1.1.28 as a whole (Bayes factor, BF=5.25).

**Fig. 2.**
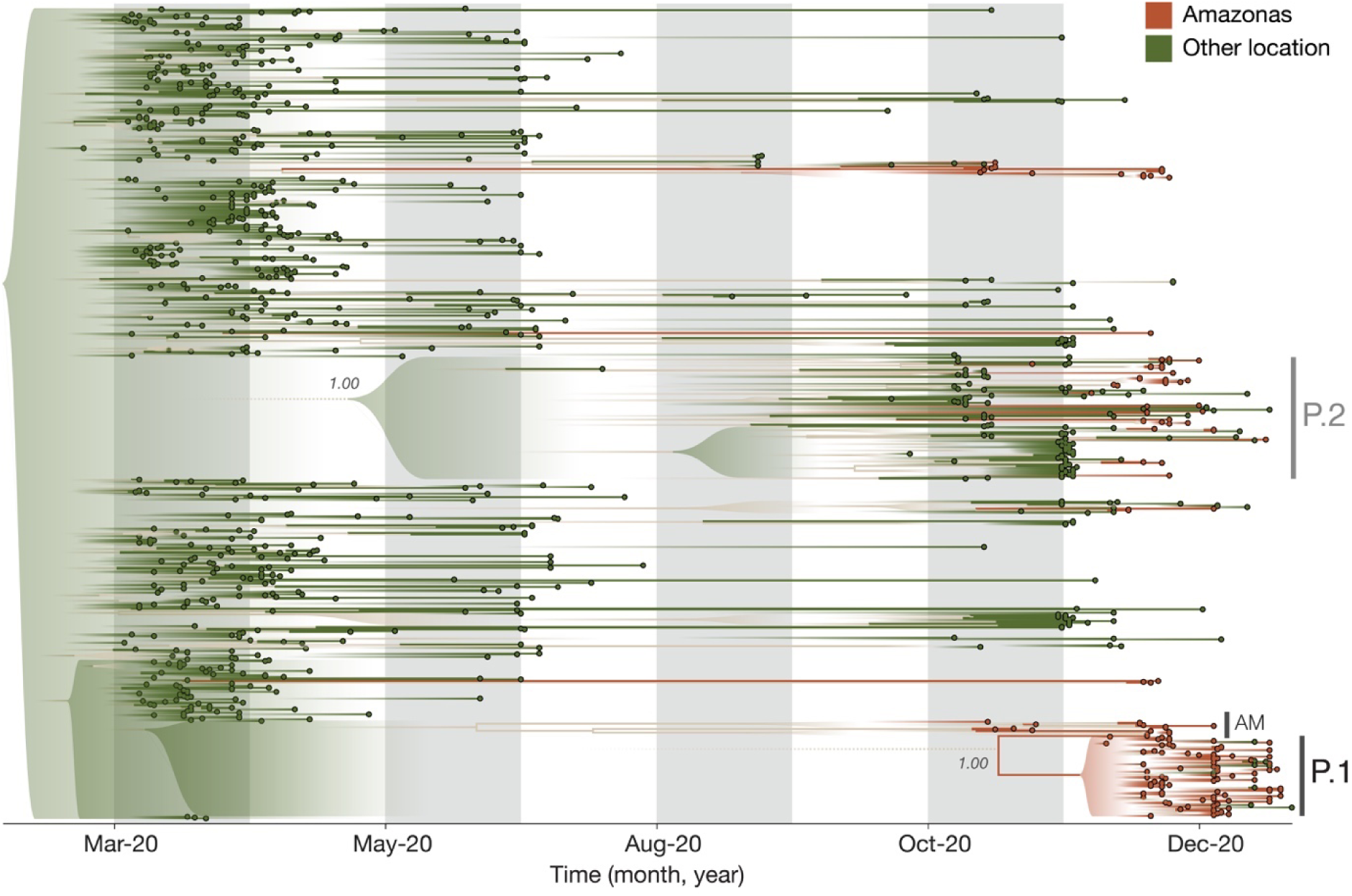
Visualization of the time-calibrated maximum clade credibility tree reconstruction for B.1.1.28, P.1 and P.2 lineages (*n*=974) in Brazil. Terminal branches and tips of Amazonas state are coloured in brown and those from other locations are coloured in green. Nodes with posterior probabilities of <0.5 have been collapsed into polytomies and their range of divergence dates are illustrated as shaded expanses.

Our data suggests multiple introductions of the P.1 lineage from Amazonas to Brazil’s south-eastern states (Fig. 2). We also detected 7 small well-supported clusters of P.2 sequences from Amazonas (2–7, posterior probability=1.00). Virus exchange between Amazonas state and the urban metropolises in southeast Brazil largely follow patterns in national air travel mobility (Fig. 1D, fig. S12).

## Infection with P.1 and sample viral loads

We analysed all SARS-CoV-2 RT-qPCR positive results from a laboratory providing testing in Manaus since May 2020 (Fig. 1A, **Data S2**) with the aim of exploring trends in sample RT-qPCR cycle threshold (Ct) values, which are inversely related to sample virus loads and transmissibility (*32*). By focusing on data from a single laboratory, we reduce instrument and process variation that can affect Ct measurements.

We analysed a set of RT-qPCR positive cases for which virus genome sequencing and lineage classification had been undertaken (*n* = 147). Using a logistic function (Fig. 3A) we find that the fraction of samples classified as P.1 increased from 0% to 87% in around 7 weeks (table S1), quantifying the trend shown in Fig 1C. We found a small but statistically significant association between P.1 infection and lower Ct values, for both the *E* gene (lognormal regression, p = 0.029, *n* = 128 samples, 61 of which were P.1) and *N* gene (p = 0.01, *n* = 129, 65 of which were P.1), with Ct values lowered by 1.43 (0.17-2.60 95% CI) and 1.91 (0.49-3.23) cycles in the P.1 lineage on average, respectively (Fig. 3B).

**Fig. 3.**
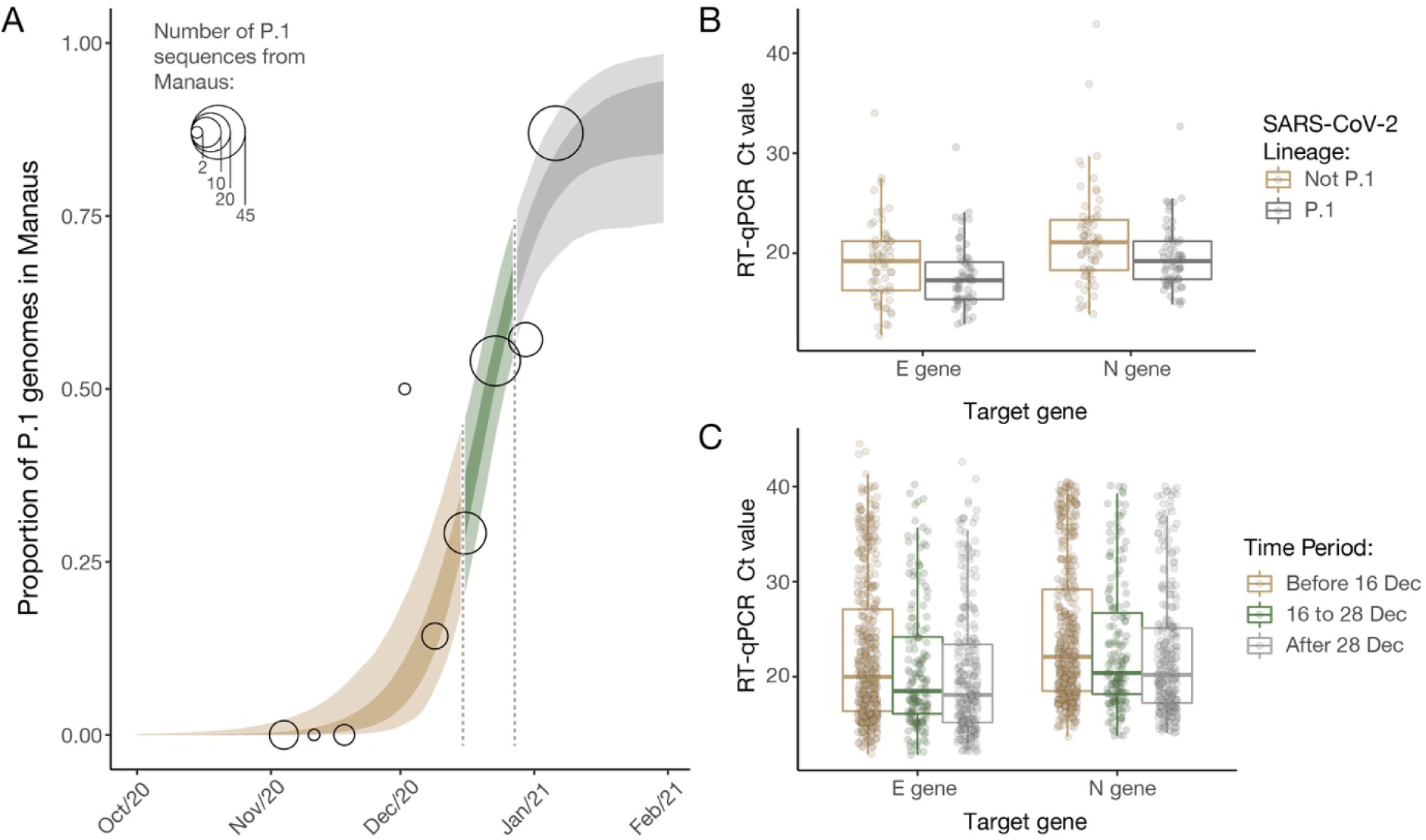
Temporal variation in the proportion of sequenced genomes belonging to P.1, and trends in RT-qPCR Ct values for COVID-19 infections in Manaus. **(A)** Logistic function fitting to the proportion of genomes in sequenced infections that have been classified as P.1 (black circles, size indicating number of infections sequenced), divided up into time-periods where the predicted proportion of infections that are due to P.1 is <1/3 (light brown), between 1/3 and 2/3 (green) and greater than 2/3 (grey). For the model fit, darker ribbon represents the 50% credible interval, and lighter ribbon represents the 95% credible interval. For the data points, grey thick line is the 50% exact Binomial confidence interval and the thinner line is the 95% exact Binomial confidence interval. **(B)** Ct values for genes *E* and *N* in a sample of symptomatic cases presenting for testing at a healthcare facility in Manaus, stratified according to the period defined in **(A)** in which the oropharyngeal and nasal swab collections occurred in. **(C)** Ct values for genes *E* and *N* in a subsample of 184 infections included in **(B)** that had their genomes sequenced (dataset A).

Using a larger sample of 942 Ct values (including an additional 795 samples for which no lineage information was available) we investigated Ct values across three time periods characterised by increasing P.1 relative abundance. Average Ct-values for both the *E* and *N* genes decline through time, as both case numbers and the fraction of P.1 infections increased (Fig. 3C; *E* gene p = 0.022 and p<0.0001 for comparison of time periods 2 and 3 to period 1; N gene p = 0.025 and p<0.001, respectively).

However, population-level Ct distributions are sensitive to changes in the average time since infection when samples are taken, such that median Ct values can decrease during epidemic growth periods and increase during epidemic decline (*33*). In an attempt to account for this effect, we assessed the association between P.1 infection and Ct levels whilst controlling for the delay between symptom onset and sample collection. Statistical significance was lost for both data sets (E gene p = 0.15, n = 42, 22 of which were P.1; N gene p = 0.12, n = 42, 22 of which were P.1). Due to this confounding factor we conclude that we cannot yet determine if P.1 infection is associated with increased viral loads (*34*) or a longer duration of infection (*35*).

## Mathematical modelling of lineage P.1 epidemiological characteristics

We next explored epidemiological scenarios that might explain the recent resurgence of transmission in Manaus (*36*). To do this, we extend a semi-mechanistic Bayesian model of SARS-CoV-2 transmissibility and mortality (*41-43*) to include two categories of virus (“P.1” and “non-P.1”) and to allow characteristics such as infection severity, transmissibility and propensity for re-infection to vary between the categories. It also integrates information on the timing of P.1 emergence in Manaus, using our molecular clock results (Fig. 2). The model explicitly incorporates waning of immune protection following infection, parameterized using dynamics from a longitudinal cohort study (*37*), to explore the competing hypothesis that waning of prior immunity might explain the observed resurgence (*36*). We use the model to evaluate the statistical support that P.1 possesses altered epidemiological characteristics compared to local non-P.1 lineages. The model is fitted to both COVID-19 mortality data (with a correction for systematic reporting delays (*38, 39*)) and to the estimated increase through time in the proportion of infections due to P.1 derived from genomic data (Table S1). We assume within-category immunity wanes over time (50% wane within a year, though sensitivity analyses are presented in Table S4) and that cross-immunity (the degree to which previous infection with a virus belonging to one category protects against subsequent infection with the other) is symmetric between categories (see details in **Supplementary Materials**).

Our results suggest the epidemiological characteristics of P.1 are different to those of previously circulating local SARS-CoV-2 lineages, but also highlight substantial uncertainty in the extent and nature of this difference. Plausible values of transmissibility and cross immunity exist in a limited area but are correlated (Fig. 4A, with the extent of immune evasion defined as 1 minus the inferred cross-immunity). This is expected, because in the model a higher degree of cross-immunity means that greater transmissibility of P.1 is required to generate a second epidemic. Within this plausible region of parameter space, P.1 can be between 1.4–2.2 (50% BCI, with a 96% posterior probability of being >1) times more transmissible than local non-P1 lineages and can evade 25-61% (50% BCI, with a 95% posterior probability of being able to evade at least 12%) of protective immunity elicited by previous infection with non-P.1 lineages (Fig. 4A). The joint-posterior distribution is inconsistent with a combination of high increased transmissibility and low cross-immunity (Fig. 4A). Moreover, our results further show that natural immunity waning alone is unlikely to explain the observed dynamics in Manaus, with support for P.1 possessing altered epidemiological characteristics robust to a range of values assumed for the date of the lineage’s emergence and the rate of natural immunity waning (Tables S3 and S4). We caution that these results are not generalisable to other settings; more detailed and direct data are needed to identify the exact degree and nature of the changes to P.1’s epidemiological characteristics compared to previously circulating lineages.

**Fig. 4.**
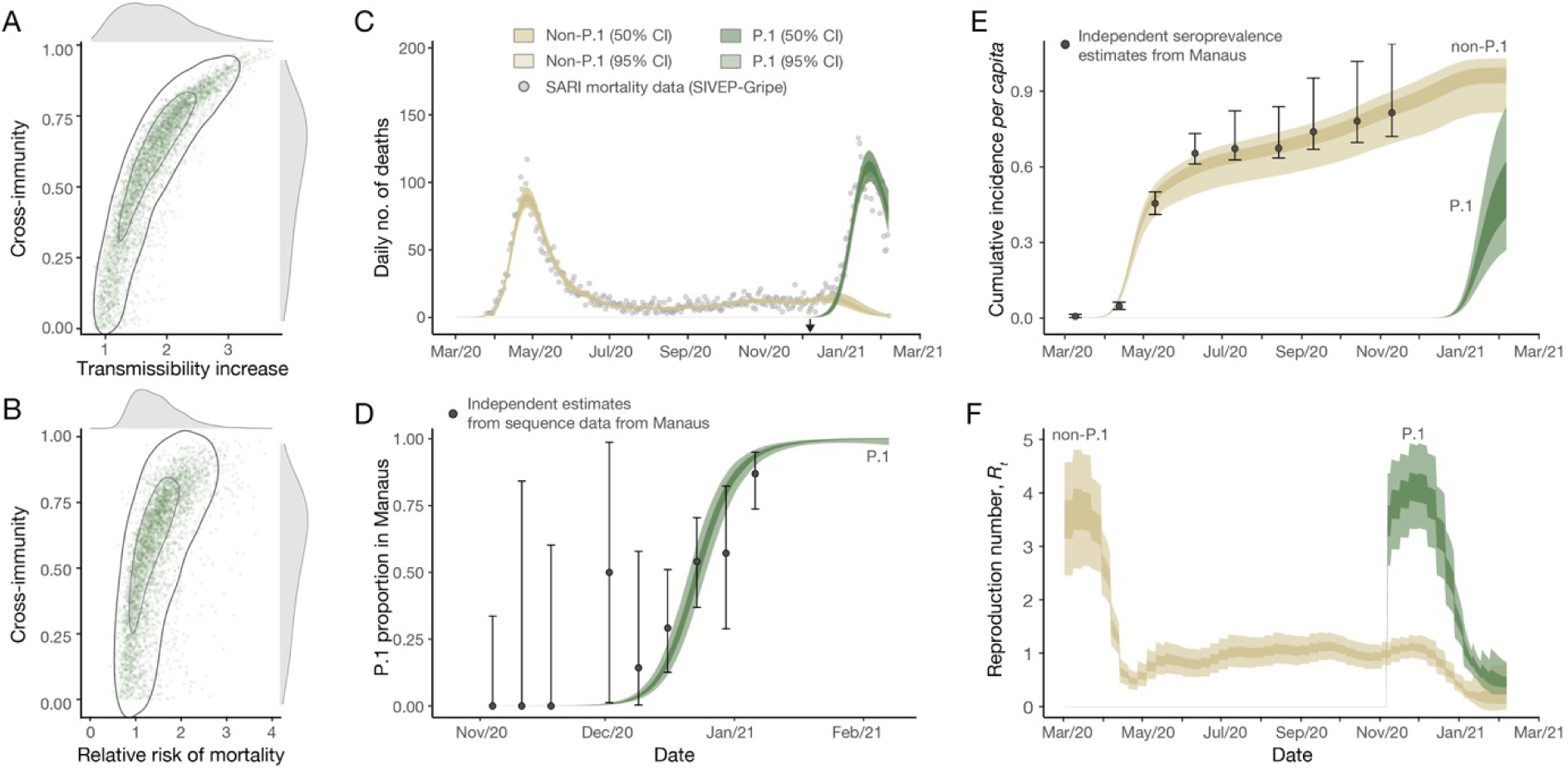
Estimates of P.1’s epidemiological characteristics inferred from a multicategory Bayesian transmission model fitted to data from Manaus, Brazil. **(A)** Joint posterior distribution of the cross-immunity and transmissibility increase inferred through fitting the model to mortality and genomic data. Grey contours refer to posterior density intervals ranging from the 95% and 50% isoclines. Marginal posterior distributions for each parameter shown along each axis. **(B)** As for (A) but showing the joint-posterior distribution of cross-immunity and the inferred relative risk of morality in the period following P.1’s emergence compared to the period prior. **(C)** Daily incidence of COVID-19 mortality. Points show severe acute respiratory mortality records from the SIVEP-Gripe database (*71*), brown and green ribbons show model fit for COVID-19 mortality incidence, disaggregated by mortality attributable to non-P.1 lineages (brown) and the P.1 lineage (green). **(D)** Estimate of the proportion of P.1 infections through time in Manaus. Black data points with error bars are the empirical proportion observed in genomically sequenced cases (see Fig. 3A) and green ribbons (dark = 50% BCI, light = 95% BCI) the model fit to the data. **(E)** Estimated cumulative infection incidence for the P.1 and non-P.1 categories. Black data points with error bars are reversion-corrected estimates of seroprevalence from blood donors in Manaus (*2*), coloured ribbons are the model predictions of cumulative infection incidence for non-P.1 lineages (brown) and P.1 lineages (green). These points are shown for reference only and were not used to fit the model. **(F)** Bayesian posterior estimates of trends in reproduction number *R_t_* for the P.1 and non-P.1 categories.

We estimate that infections are 1.1–1.8 (50% BCI, 81% posterior probability of being >1) times more likely to result in mortality in the period following P.1’s emergence, compared to before, although posterior estimates of this relative risk are also correlated with inferred cross-immunity (Fig. 4B). More broadly, the recent epidemic in Manaus has strained the city’s healthcare system leading to inadequate access to medical care (*40*). We therefore cannot determine whether the estimated increase in relative mortality risk is due to P.1 infection, stresses on the Manaus healthcare system, or both. Detailed clinical investigations of P.1 infections are needed. The model fits well observed time series data from Manaus on COVID-19 mortality (Fig. 4C) and the relative frequency of P.1 infections (Fig. 4D) and also captures previously-estimated trends in cumulative seropositivity in the city (Fig. 4E). We estimate the reproduction number (*Rt*) on 07 Feb 2021 to be 0.2 (50% BCI: 0.1-0.3) for non-P.1 and 0.5 (50% BCI: 0.4-0.6) for P.1 (Fig. 4F).

## Characterisation and adaptation of a constellation of spike protein mutations

Lineage P.1 contains 10 new amino acid mutations in the virus spike protein (L18F, T20N, P26S, D138Y, R190S, K417T, E484K, N501Y, H655Y, T1027I) compared its immediate ancestor (B.1.1.28). In addition to the abovementioned estimated increase in the rate of molecular evolution during the emergence of P.1, we find, using molecular selection analyses (*41*), evidence that 9 of these 10 mutations are under diversifying positive selection (Fig. S11).

Three key mutations present in P.1, N501Y, K417T and E484K, are located in the spike protein RBD. The former two interact with human angiotensin-converting enzyme 2 (hACE2) (*11*), whilst E484K is located in a loop region outside the direct hACE2 interface (fig. S11). Notably, the same three residues are mutated with the B.1.351 variant of concern, and N501Y is also present in the B.1.1.7 lineage. The independent emergence of the same constellation of mutations in geographically-distinct lineages suggests a process of convergent molecular adaptation. Similar to what was observed for SARS-CoV-1 (*42-44*), mutations in the RBD may increase affinity of the virus for host ACE2 and consequently impact host cell entry and virus transmission. Recent molecular analysis of B.1.351 (*45*) suggests that the three P.1 RBD mutations may similarly enhance hACE2 engagement, providing a plausible hypothesis for an increase in transmissibility of the P.1 lineage. Moreover, E484K has been associated with reduced antibody neutralisation (*6, 46-48*) and as RBD-presented epitopes account for ∼90% of the neutralising activity of sera from individuals previously infected with SARS-CoV-2 (*49*), tighter binding of P.1 viruses to hACE2 may further reduce the effectiveness of neutralizing antibodies that are competing with hACE2 to bind the RBD. However, it remains difficult to estimate the contributions of the P.1 RBD mutations to transmissibility and neutralisation that may have led to P.1’s emergence at the population level.

## Conclusion

Genomic surveillance and early data sharing by teams worldwide led to the rapid detection and characterisation of P.1 (*23*), yet such surveillance is still limited in many settings. Existing genomic surveillance is currently inadequate to determine the true international extent of P.1, and this paucity limits the detection of similar variants of concern globally. Studies to evaluate real-world vaccine efficacy in response to P.1 are urgently needed, although we note that neutralisation titres represent only one component of the elicited response to vaccines, and that minimal reduction of neutralisation titres relative to earlier circulating strains is not uncommon (*45*). Until an equitable allocation and access to effective vaccines is available to all, non-pharmaceutical interventions should continue to play an important role in reducing the emergence of new variants.

## Data Availability

Preliminary genome sequences generated from samples obtained from laboratory A were shared on GISAID on 12 January 2021. Findings were shared with representatives from the World Health Organization, Pan American Health Organization, Secretary of Health Amazonas, and FioCruz Manaus on 11 January 2021. Preliminary report describing first P.1 genomes from Manaus was shared on 12 January 2021 (86). Epidemiological data and epidemiological model code, together with BEAST XML files, tree files, log files are archived at https://github.com/CADDE-CENTRE. GISAID IDs for the SARS-CoV-2 Manaus sequence data can be found in Data S2. All consensus sequences generated by this study can be found at https://github.com/CADDE-CENTRE.

## Acknowledgments

**General**: We thank Lucy Matkin (University of Oxford) for logistic support and Claudio Sachi (Instituto Adolfo Lutz) for agreeing with the use of unpublished sequence data available in GISAID before publication. We thank the administrators of the GISAID database for supporting rapid and transparent sharing of genomic data during the COVID-19 pandemic. A full list acknowledging the authors publishing data used in this study can be found in **Data S3**.

## Author contributions

Conceptualization: NRF, TAM, CW, ICM, DSC, AR, CD, OGP, SF, SB, ECS Methodology: NRF, TAM, CW, TAM, ICM, DSC, SM, FCS, IH, MSR, JGJ, LAMF, PSA, TMC, CAMS, ERM, JTM, RHM, PSP, MUK, RH, TB, OGP, MS, SP, OR, NF, NJL, PL, AR, CD, OGP, SF, SM, ECS Investigation: NRF, TAM, CW, IMC, DSC, SM, MAEC, FCSS, IH, MSR, JGJ, LAMF, PSA, TMC, CAMS, ERM, JTM, RHMP, PSP, MUK, RH, NG, WMS, LJTA, CCC, HH, GMF, ECR, LMS, MCP, FSVM, ABL, JPS, DAGZ, ACSF, RPS, DJL, PGTW, HS, ALPS, MSV, CCC, VSDC, RMFF, HMS, RSA,, BN, JH, MM, XM, HC, RS, AG, MS, TB, SP, CHW, OR, NMF, NJL, PL, AR, NAF, MPSSC, CD, OGP, SF, SB, ECS Visualization: NRF, TAM, CW, DSC, IMC, JT, AR, SP, TB, CW, SB Funding acquisition: NRF, NL, AR, OGP, NF, SF, SB, ECS Project administration: NRF, ECS Supervision: NRF, OGP, AR, CD, NL, SB, ECS Writing – original draft: NRF, TM, CW, IMC, DSC, SF, SB, OGP, CD, ECS Writing – review & editing: All authors

## Data and materials availability

All data, code, and materials used in the analysis are available in a dedicated GitHub Repository: https://github.com/CADDE-CENTRE.

## Supplementary Materials

### Materials and Methods

#### Ethics

Residual oropharyngeal and nasal swab collections from Manaus residents testing positive for SARS-CoV-2 RT-qPCR between 1 November 2020 and 9 January 2021 were obtained from two private clinical laboratories in Manaus. All samples were de-identified before receipt by the researchers. Ethical approval for this study was confirmed by the national ethical review board (Comissão Nacional de Ética em Pesquisa), protocol number CAAE 30127020.0.0000.0068.

#### Sampling and Metadata Collection

A total of 436 SARS-CoV-2 samples RT-qPCR confirmed or suspected were collected for genomic sequencing between 1 November 2020 and 9 January 2021. Samples were provided for confirmatory testing and genome sequencing. For clinical laboratory A (*n*=37 RT-qPCR positive cases with sampling dates from 15 to 23 December 2020), SARS-CoV-2 diagnosis was performed using the Allplex 2019-nCoV Assay (Seegene, South Korea) assay that detects *RdRP*, *N* specific genes for SARS-CoV-2 and the *E* gene for all Sarbecovirus subgenus, including SARS-CoV-2 (*50, 51*). For clinical laboratory B, 399 RT-qPCR positive or suspected cases [representing 73% of all 548 samples with a RT-qPCR positive (*n*=545) or inconclusive (*n*=3) results between 2 November and 9 January 2021] were processed for genome sequencing. In this case, RT-qPCR was determined using the Xpert Xpress SARS-CoV-2 platform (GeneXpert) that detects the nucleocapsid (*N*) gene specific for SARS-CoV-2 and the envelop (*E*) gene of *Sarbecovirus* subgenus including SARS-CoV-2 (Cepheid, USA). Samples were shipped in dry ice to the Institute of Tropical Medicine, University of São Paulo, Brazil, for genome sequencing. Also, RT-qPCR cycle threshold values and associated metadata (patient age and sex, date of onset symptom, date of RT-qPCR test, data of sample collection when available, cycle threshold, Ct, values for *E* and *N* genes) were recorded for 1,084 RT-qPCR positive and 16 inconclusive results from laboratory B in Manaus between 18 May 2020 and 27 January 2021 (**Data S1**).

#### PCR Amplification and Virus Nanopore Sequencing

Viral RNA was isolated from 200-μl SARS-CoV-2-suspected samples using the QIAamp Viral RNA Mini kit (QIAGEN, Hilden, Germany) according to the manufacturer’s instructions. Virus genome sequencing was carried out on all positive samples regardless of laboratory reported RT-qPCR cycle threshold values using a combination of targeted multiplex-PCR amplification and portable nanopore sequencing MinION platform (Oxford Nanopore Technologies, ONT, UK).

cDNA synthesis was performed from the extracted RNA using random hexamers, and the Protoscript II First Strand cDNA synthesis Kit (New England Biolabs, UK). Subsequently, the ARTIC network SARS-CoV-2 V3 primer scheme and Q5 High-Fidelity DNA polymerase (New England Biolabs, UK) were used for SARS-CoV-2 whole-genome multiplex-PCR amplification (*22*). AmpureXP beads (Beckman Coulter, United Kingdom) were used for PCR product purification and fluorimetry-based quantification was carried out using the Qubit dsDNA High Sensitivity assay on the Qubit 3.0 (Life Technologies, USA). To ensure uniform sequencing of samples, equimolar normalisation of 10 ng per sample was performed followed by barcoding using the EXP-NBD104 (1–12) and EXP-NBD114 (13–24) Native Barcoding Kits (Oxford Nanopore Technologies, UK). Finally, barcoded samples were pooled followed by library preparation using the SQK-LSK109 Kit (Oxford Nanopore Technologies, UK).

Nanopore sequencing libraries were loaded onto an R9.4.1 flow-cell (Oxford Nanopore Technologies, UK) and sequenced using MinKNOW version 20.10.3 (Oxford Nanopore Technologies, UK). FAST5 files containing the raw signal data were basecalled, demultiplexed, and trimmed using Guppy v4.4.1 (Oxford Nanopore Technologies, UK). The process reads were aligned against the reference genome Wuhan-Hu-1 (GenBank: MN908947.3) using minimap2 v2.17.r941 and converted to a sorted BAM file using SAMtools (*52*). Length filtering, quality test and primmer trimming was performed for each barcode using artic guppyplex and variant calling and consensus sequences using artic minion with Nanopolish and Medaka versions from ARTIC bioinformatics pipeline ( https://github.com/artic-network/fieldbioinformatics). Genome regions with a depth of <20-fold were represented with N characters. The genome statistics were obtained from SAMtools and the Tablet viewer (*53*). Any runs suspected to have any level of contamination were discarded. Individual nanopore sequencing statistics for each sequence can be found in **Data S2**.

#### Genome Datasets

Genome coverage of 184 generated sequences obtained from clinical samples varied from 27 to 99% of the virus SARS-CoV-2 genome. Of these, 35 sequences (average Ct of 21.78, range 14.5–29.2) had a virus genome coverage between 25–75%. However, even partial sequences can provide important information about changes in SARS-CoV-2 lineage structure (*53*).

We compiled three genome datasets from Manaus from data generated in this study: *dataset A* included 184 sequences with >25% virus genome coverage (37 from laboratory A and 147 from laboratory B) and was used to estimate virus lineage frequency in Manaus over time; *dataset B* included 143 near-complete genome sequences with >75% of the virus genome coverage (31 from laboratory A, 112 from laboratory B); and *dataset C* included 48 sequences with >95% of the virus genome complete (*n*=17 from laboratory A, *n*=31 from laboratory B).

For *datasets A, B and C*, a reference genome sequence Wuhan-Hu-1 (GenBank: MN908947.3) was appended before multiple sequence alignment using MAFFTv.7 (*54*). For *dataset A*, lineage classification was conducted using manual phylogenetic analysis. Sequences with genome coverage between 25% and 75% were appended to *dataset C* and assigned to B.1.1.28, P.2 and P.1 lineages based on monophyletic clustering of each sequence within each of these lineages (fig. S2). Manual phylogenetic subtyping and PANGO lineage classification using the latest pangolin version (v.2.2.1, 6 February 2021) (*15*) was conducted for *dataset B* and *dataset C* (figs. S3-S5). Date of sample collection, age, sex, RT-qPCR CT values, lineage assignment and sequencing statistics for 184 sequences generated in this study from Manaus can be found in **Data S2**.

We downloaded all sequences publicly available in GISAID (up to 14-01-2021) and selected for analysis those that were published in *PubMed*, *MedRxiv*, *BioRxiv* or *Preprint* repositories. Specifically, *dataset B* and *dataset C* from Manaus were appended to (*i*) B1.1.28 genome sequences with >95% virus genome coverage from the Brazilian Amazon (*55*), Minas Gerais (*56*), Pernambuco (*57*), Rio de Janeiro (*58*, *59*), Rio Grande do Sul (*60*) and to data from an early country-wide study of SARS-CoV-2 diversity in Brazil (*17*). *Datasets B* and *C* were also appended to (*ii*) P.2 genome sequences with >95% virus genome coverage from Rio de Janeiro (*59*), Rio Grande do Sul (*60*, *61*). Exceptionally, written permission was obtained from the reference laboratory in São Paulo, Institute Adolfo Lutz, to use P.1 complete genome sequences shared in GISAID (up to 19-01-2021) as part of their surveillance activities. Duplicate sequences were removed from the alignments, 5’ and 3’ untranslated regions from each genome were discarded. A table describing GISAID IDs, authors, originating and submitting laboratory for all publicly available data used in *dataset B’* and *dataset C’* (with data from this study and publicly available data) can be found in **Data S3**.

#### Maximum Likelihood Tree Reconstruction

Fast and efficient maximum likelihood (ML) phylogenetic trees were reconstructed using IQTREE 2 (*62*) for *dataset A’* (*n*=999) (used for lineage classification of >25 and <75% virus genome coverage samples from Manaus, **Data S2**), and for the B.1.28, P.1 and P.2-specific *dataset B’* (*n*=974) and *dataset C’* (*n*=883) (ML trees can be found in figs. S2-S4). Briefly, a Jukes Cantor (*63*) DNA substitution model assuming equal substitution rates and equal base frequencies. Near zero branches were collapsed so that the final tree could be multifurcating to account for the many polytomies observed in SARS-CoV-2 phylogenetic trees. To explore temporal structure of *dataset B’* and *dataset C’*, root-to-tip genetic distances (*d*) were regressed against sampling dates (yyyy-mm-dd) using TempEst v.1.5.3 (*64*). One sequence showed incongruent genetic diversity compared with its sampling date (SP-322 EPI-ISL-693197) and was discarded from subsequent analyses. Strong and identical correlations between *d* and sequence sampling dates were observed for *dataset B’* (r^2^=0.80) (fig. S6) and *dataset C’* (r^2^=0.80) (fig. S7). Therefore, we used *dataset B’* for subsequent phylogenetic analyses. Regressions between root-to-tip distances and sampling dates in *dataset B’* were also fit separately for B.1.1.28, P.2, and P.1 lineages in R v.3.6.2 (*54*). These showed no obvious difference in evolutionary rates, and suggested that P.2 and P.1 show a tendency to fall over the regression line of B.1.1.28 (fig. S8). This supports an increased evolutionary rate on the ancestral branches leading to P.1 and P.2 compared to B.1.1.28 evolutionary rate, an evolutionary scenario that seems to be characteristic of SARS-CoV-2 lineages of concern (*13*).

#### Bayesian Coalescent Inference

Next, we used a fully probabilistic Bayesian framework to reconstruct molecular clock phylogenies and estimate growth rates directly from time-stamped genome sequence data. Substitution rates were modelled according to a HKY with 4 gamma categories to account for among site rate variation (*65*, *66*). The B.1.1.28 lineage has been circulating in Brazil since late February-early March (*17*). P.1 and P.2 lineages are phylogenetically nested within the more diverse and older B.1.1.28 strains and form separate monophyletic clades (see figs. S2-S5).

To account for rate differences in ancestral branches leading to P.1 and P.2, we estimate molecular clock trees using a local clock implementation in a Bayesian framework (*69*) that explicitly allows for distinct evolutionary rates on the ancestral branch leading to P.1 and P.2. Local clock models allow different rates along distinct lineages of a single phylogeny (*69*), which may be particularly suitable to estimate dates of emergence of P.1 and P.2 because they can take into account higher rates of mutation accumulation over short periods of time that could to be linked to selective pressures associated with the emergence of lineages of concern (*70*). Analyses were run with a flexible non-parametric skygrid tree prior (*31*) in triplicate for 100 million MCMC steps using BEAST version 1.10.4 (*55*) with BEAGLE library v3.1.0 for accelerated likelihood evaluation (*56*). Parameters and trees were sampled every 10,000 steps and convergence of MCMC chains was inspected with Tracer v1.7.1 (*57*). Posterior probability distributions for the most recent common ancestor of the P.1 and P.2 clade are shown in fig. S9.

We also used a nested coalescent model (*58*) to estimate viral growth rates for B.1.1.28, and P.2. We fit a constant demographic model to the phylogeny excluding the P.1 and P.2 clades. Two separate logistic growth models were then fitted to P.1 and P.2 clades. For the logistic growth coalescent models, a lognormal prior with mean 1.0 and a standard deviation of 10 were used for the population size; for the growth rate, we used a Laplace prior with a mean of 0 and a scale of 10. For the local clock model, we use a normal prior with mean −7.0 and standard deviation of 5 on the background rate in log space and a normal prior with mean 0 and standard deviation of 0.5 for the log effect sizes on the branches for which the rates are allowed to deviate from the background rate (*59*). All other priors used for phylogenetic inference were kept at default settings. Posterior probability distributions for the most recent common ancestor of the P.1 and P.2 clade according to the constant-logistic-logistic model and corresponding growth rate and doubling time parameters are show as fig. S9 and fig. S10.

To quantify the support for both the rate differences in the local clock model and the growth rates in the nested coalescent model, we conduct Bayes Factor (BF) tests. For this purpose, we employ posterior indicator functions that allow estimating the posterior probability that a specific substitution rate (the rate on the branch ancestral to P.1 or P.2) is larger than another rate (the background rate for B.1.1.28) and that one growth rate (for P.1) is larger than another growth rate (for P.2). We use these posterior probabilities to calculate the BF as the ratio of the posterior odds over the prior odds that substitution rates or growth rates are different, assuming that the prior probabilities for these differences are 0.5 (in line with our prior specification on these parameters).

#### Adaptive Evolution of P.1 lineage

We investigated the extent of selective forces acting on P.1 and P.2 SARS-CoV-2 lineages using HyPhy v2.5.27 (*77*). We analyzed a median of 5 unique P.1 haplotypes per gene/peptide in the context of a median of 79 reference sequences, and a median of 9 unique P.2 haplotypes per gene/peptide in the context of a median of 70 reference sequences. The summary of sites subject to episodic diversifying selection (p≤0.05) identified using MEME (*60*) is shown in Table S2. In addition to individual sites under selection, we also recorded instances of putative convergence, i.e., substitutions to the same amino-acid at the same site in both lineages; there were only 2 such events (S/484K and ORF1A/318L). The evolutionary “credibility” of target residues was estimated using the <u>PRIME method</u> (*77*) based on a <u>bat/pangolin Sarbecovirus alignment (*61*)</u>. Results can be visualized at https://hackmd.io/7hFvRdJdSVSONv_wW40_Rg.

#### Structural Analysis of P.1 lineage

Lineage defining mutations were mapped onto a previously reported cryoEM structure of the cleaved trimeric SARS-CoV-2 S ectodomain [PDB: 6ZGI, (*62*)] with PyMOL v 2.4.0 (*63*) (fig. S11). Substituted residues are indicated as spheres and coloured by type of selection according to MEME support (Table S2). Similarly, SARS-CoV-2 S RBD-hACE2 contact residues were mapped as observed in the RBD-hACE2 complex crystal structure [PDB: 6MOJ, (*11*)] together with RBD-resident substitutions specific to the P.1 lineage (fig. S11). N-linked glycans are omitted for clarity.

#### Air Travel and Mobile Geolocation Data

To better contextualize the spread of P.1 lineage within Brazil, we investigate two different mobility data sources. First, we analyse monthly air passenger travel data produced by Brazil’s Civil Aviation Agency (ANAC) which is publicly available at https://www.anac.gov.br/assuntos/setorregulado/empresas/envio-de-informacoes/base-de-dados-estatisticos-do-transporte-aereo. This includes the number of passengers and connections for international flights to and from Brazil, as well as domestic flights within the country. Using this data, we calculated the total number of passengers who travelled from Manaus between November and December 2020 disaggregated by state of origin and destination (Fig. 1D, fig. S12).

State-level mobility where the origin of the trip was the municipality of Manaus were calculated from approximately 5 million trips aggregated from anonymized cell phone data users in the month of November 2020 (*85*) (fig. S12). Data was obtained from In Loco (<u>mapabrasileirodacovid.inloco.com.br</u>), a company that provides geolocation services for a broad range of mobile applications and covers ∼20% of the mobile devices in the country. Anonymized cell-phone shows a similar pattern to air travel data, but shows travel from Manaus to other municipalities in the Amazonas states being even more important than with flights. Numbers of recorded state-level movements from and to Amazonas state, as well as city-level movements from and to Manaus municipality, are available as **Data S5** and **Data S6**, respectively.

#### Logistic Function Fitting to P.1 Genome Fraction

We fitted a logistic function to the time-varying fraction of sequenced genomes belonging to P.1, binned according to the week sampling had occurred in. The form of the logistic function is as follows:

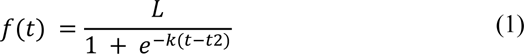

where *L* = maximum value of the logistic function, *k* = logistic growth rate, *t* = time since P.1’s emergence and *t*2 is the (inferred) time at which half of the genomes in sequenced cases belong to P.1. Model fitting was carried out using a Bayesian framework, written in the probabilistic programming language STAN and implemented in the statistical software R (Version 4.0.2) using the package *rStan* (Version 2.19.03). Two chains of 1000 iterations each were run, with the 1st 500 samples from each discarded as burn-in and the remaining 500 (for each chain, 1000 samples total) retained for inference.

#### Description of the Epidemiological Model

We utilise a Bayesian semi-mechanistic model of SARS-CoV-2 spread and mortality based on a renewal-process equation (*33*, *34*, *84*) and extended to include i) multiple SARS-CoV-2 lineages introduced at different points in time; ii) the possibility for these different lineages to possess distinct epidemiological characteristics (such as severity, transmissibility and immune evasion); and iii) waning of natural immunity due to prior infection - parameterised from the results of the recent Public Health England SIREN study - a longitudinal cohort study tracking (re)infection in healthcare workers in the United Kingdom (*36*).

Here, we model two different SARS-CoV-2 lineages, hereafter referred to as P.1 and non-P.1, with the timing of P.1’s emergence based on the phylogenetic analyses described previously. A full mathematical description of the model and its associated parameters are available in **Supplementary Text.** We run a number of model scenarios in order to evaluate support for P.1 possessing distinct epidemiological characteristics, specifically running multiple models that vary in their assumption surrounding timing of P.1 emergence. For this, we explore a total of 5 scenarios defined according to the 95% Bayesian Credible Interval for the most recent common ancestor of the P.1 lineage, including a central scenario of the 6 November 2020, two scenarios of plus or minus one week, an earlier emergence of 9 October 2020, and a later emergence of 30 November 2020 (tables S3 and S4). We also vary our assumptions surrounding the duration of protective immunity following infection (i.e. the rate at which natural immunity elicited by prior SARS-CoV-2 infection declines).

The model is fitted to two sources of data. Mortality data from the SIVEP-Gripe (*Sistema de Informação de Vigilância Epidemiológica da Gripe*) SARI (severe acute respiratory infections) hospitalisation database (*37*), including both class 4 and 5 death records (corresponding to confirmed and suspected COVID-19 deaths), consistent with earlier analyses (*8*) and corrected for known delays in mortality reporting using a Gaussian process nowcasting based framework (*38, 39*). In addition to COVID-19 mortality data, we also integrate genomic data from the sequenced samples, fitting the model to the fraction of sequenced genomes each week that belong to P.1 described in table S1.

Model fitting was carried out using a Bayesian framework, written in the probabilistic programming language STAN and implemented in the statistical software R (Version 4.0.2) using the package *rStan* (Version 2.19.03). Hamiltonian Monte Carlo with 3 chains of 1000 iterations each were run, with half the samples discarded as burnin and the remaining retained for inference. In every instance chains mixing was satisfactory, with traditional rhat statistics (for assessing convergence) less than 1.02. All code used for inference and plotting is available at https://github.com/CADDE-CENTRE.

#### Data sharing and code availability

Preliminary genome sequences generated from samples obtained from laboratory A were shared on GISAID on 12 January 2021. Findings were shared with representatives from the World Health Organization, Pan American Health Organization, Secretary of Health Amazonas, and FioCruz Manaus on 11 January 2021. Preliminary report describing first P.1 genomes from Manaus was shared on 12 January 2021 (*86*). Epidemiological data and epidemiological model code, together with BEAST XML files, tree files, log files are archived at https://github.com/CADDE-CENTRE. GISAID IDs for the SARS-CoV-2 Manaus sequence data can be found in **Data S2**. All consensus sequences generated by this study can be found at https://github.com/CADDE-CENTRE.

#### Epidemiological Model

This work builds on a previously published mathematical model of SARS-CoV-2 transmission introduced in Flaxman et al, 2020 (*64*). Specifically, we extend this semi-mechanistic Bayesian model to include multiple SARS-CoV-2 strains and the possibility for these strains to possess distinct, strain-specific epidemiological characteristics (such as transmissibility, ability to evade prior immunity, and severity of COVID-19 disease elicited).

Although any number of strains are possible within the following framework, we consider only two strains here, defined as *s* ∈ {1,2}. For strain 1, the population-unadjusted reproduction number is defined as follows:

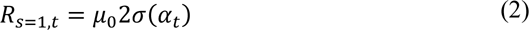

where *μ* is a scale parameter (3.3), *σ* is a logistic function, and *α* _t_ is an second-order autoregressive process with weekly time innovations. The population-unadjusted reproduction number of the second strain is modelled as:

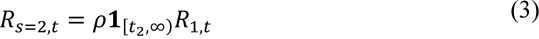

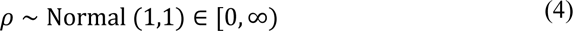

where *ρ* is a parameter defining the relative transmissibility of strain 2 compared to strain 1 and **1**_[*t*2,∞)_ is an indicator function taking the value of 0 prior to *t*_2_, and 1 thereafter, highlighting that strain 2 does not contribute to the observed evolution of the epidemic before its emergence. Introduction of the second strain at time *t*_2_ is informed through our local molecular clock analysis (see Fig. 2, Fig. S9). We note that the reproduction number estimates take into account the effect of population-level immunity and behavioural changes (modelled using a latent stochastic process). For the purposes of the primary results presented in the main text, it is assumed *t*_2_= 6 Nov 2020, though four additional scenarios are presented varying the assumed date of P.1 emergence (see tables S3 and S4).

Infections arise for each strain according to a discrete renewal process (*65, 66*):

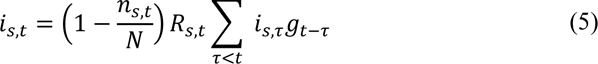

where *N* is the total population size, *n*_*s*,*t*_ is the total extent of population immunity to strain *s* present at time *t* (accounting for the cumulative number of infections with strain *s*, the extent to which immunity from these infections has waned, and the degree of cross-protection infections with other strains provide, all of which are described in more detail below). The generation of infections is then determined by the fraction of the population susceptible and available to be infected, as well as the time-varying reproduction numbers of each strain *R*_*s*,*t*_ and generation time distribution *g* from ref. (*64*).

For the original strain, infections are seeded for six days as:

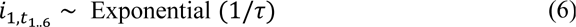

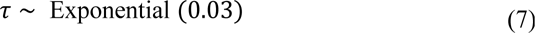

and the second strain for 1 day (*t*_2_) as:

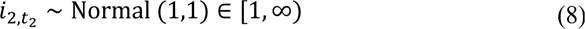

The susceptible depletion term for strain *s* is modelled as

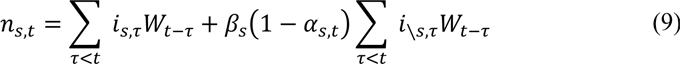

The first term describes the contribution of prior infections with strain *s* to population-level immunity for *s*. The second term describes the contribution of prior infections with not strain *s* (i. e. _\_ *s*), which is a function of the assumed cross-immunity, *β*_*s*_ *ε* [0,1]. With this formulation, β = 0 indicates no cross-protection between infections caused by different strains and β = 1 indicates complete cross-protection between infections caused by different strains. In practice we only consider symmetric cross-immunity *β*_*s*_ = *β*, which is given the prior:

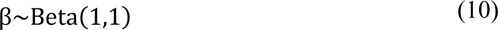

We also define *W*_*t*−*τ*_ as the time-dependent waning of immunity elicited by previous infection. This is modelled as a Rayleigh survival-type function, and is parameterized based on results from the Public Health England SIREN trial (*37*); based on these results, we use a Rayleigh parameter of *σ* = 310 which produces 50% of individuals still immune after 1 year, though explore a range of other scenarios with different assumed rates of waning (see Table S4). The cross-immunity susceptible term *α*_*s*,*t*_ is then modelled as:

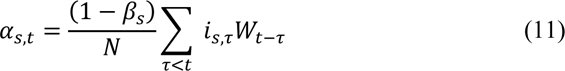

and describes the proportion of infections with variant \*s* expected to occur in individuals who have been previously infected with variant *s* - itself a function of both cross-immunity (*β*) and the proportion of the population previously infected with *s*. Infections in the model generate deaths via the following mechanistic relationship:

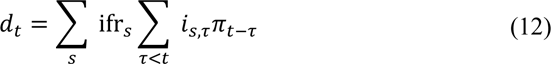

with infection fatality ratio priors:

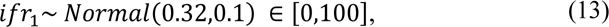

based on results from (*67*), adjusted for the age structure of the population of Manaus and:

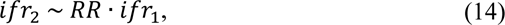

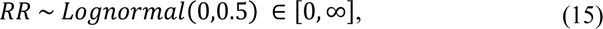

where *RR* denotes relative risk. We note that other work has shown that the degree of transmission advantage exhibited by the B.1.1.7 can vary over time as a function of control interventions and behavior (*68*).

The infection-to-death distribution *π* is composed of infection-to-onset and onset-to-death contributions as in previous work (*69*), with adaptations to take into account the most likely onset-to-death distribution in Amazonas state based on hospitalization distributions obtained by ref. (*70*).

The observation model uses two sources of data. In the first likelihood, the expected deaths *D*_*t*_ are modelled as negative-binomially distributed:

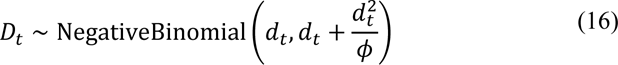

with mortality data *d*_*t*_ and dispersion prior:

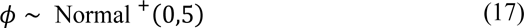

The deaths data source is based on class 4 and 5 (confirmed and probable) SARI COVID-19 deaths (*71, 72*), from the SIVEP-Gripe database for Manaus city, that have been amended with Gaussian Process nowcasting to correct for known delays between deaths occurring and being recorded in the dataset (*38, 39*).

The second likelihood is based on genomic data from symptomatic individuals presenting for testing and who had both a positive PCR diagnosis and the infecting SARS-CoV-2 genome sequenced. Specifically, the proportion of sequenced genomes identified as P.1 at time *t* are modelled with a binomial likelihood:

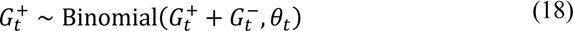

with positive counts for P.1 denoted *G*_*t*_^+^ and counts for lineages not belonging to P.1 recorded as *G*_*t*_^−^. The success probability for P.1 positivity is modelled as the infection ratio:

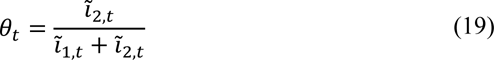

where *ĩ*_*s,t*_ is given by:

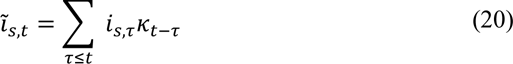

to account for the time varying PCR positivity displayed over the natural course of a COVID-19 infection. The distribution *κ* describes the probability of being PCR positive over time following infection, and is based on ref. (*73*).

For purposes of comparison with serological data, we evaluate:

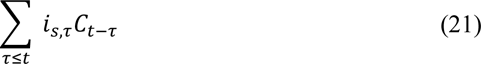

where *C*_*t*−*τ*_ is the cumulative probability of an individual infected on day τ having seroconverted by time *t*. This distribution is empirical and based on ref. (*74*).

---

**Fig. S1.**
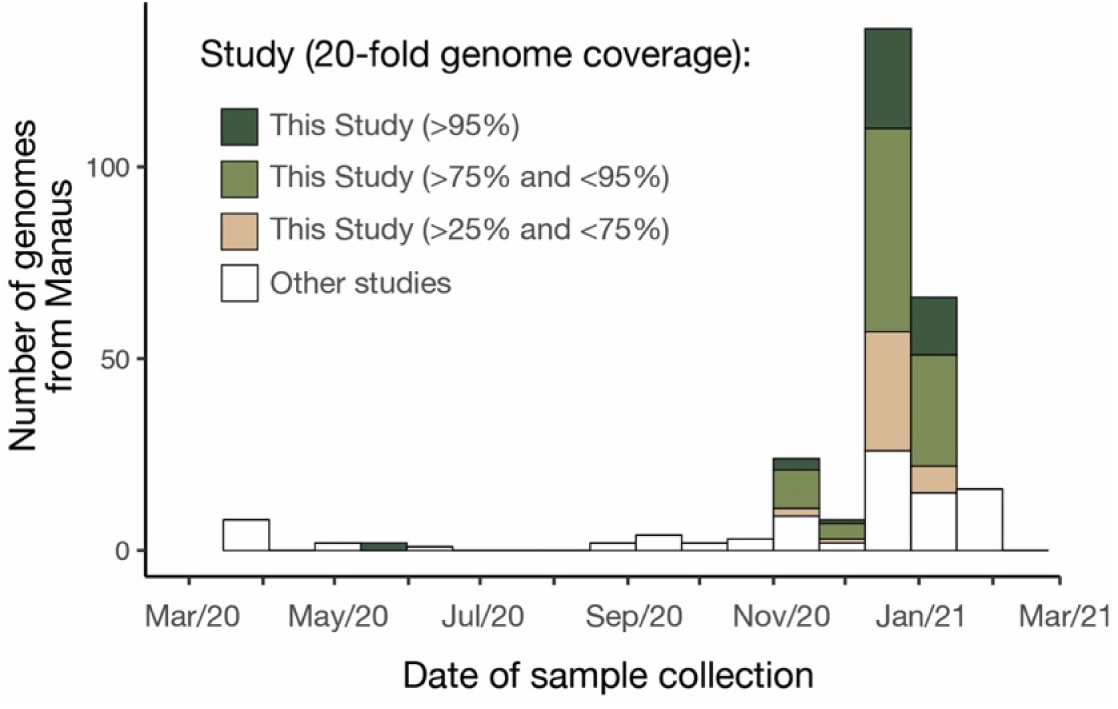
Number of genome sequences from Manaus grouped by sequence coverage and study (see also Materials and Methods).

**Figure S2.**
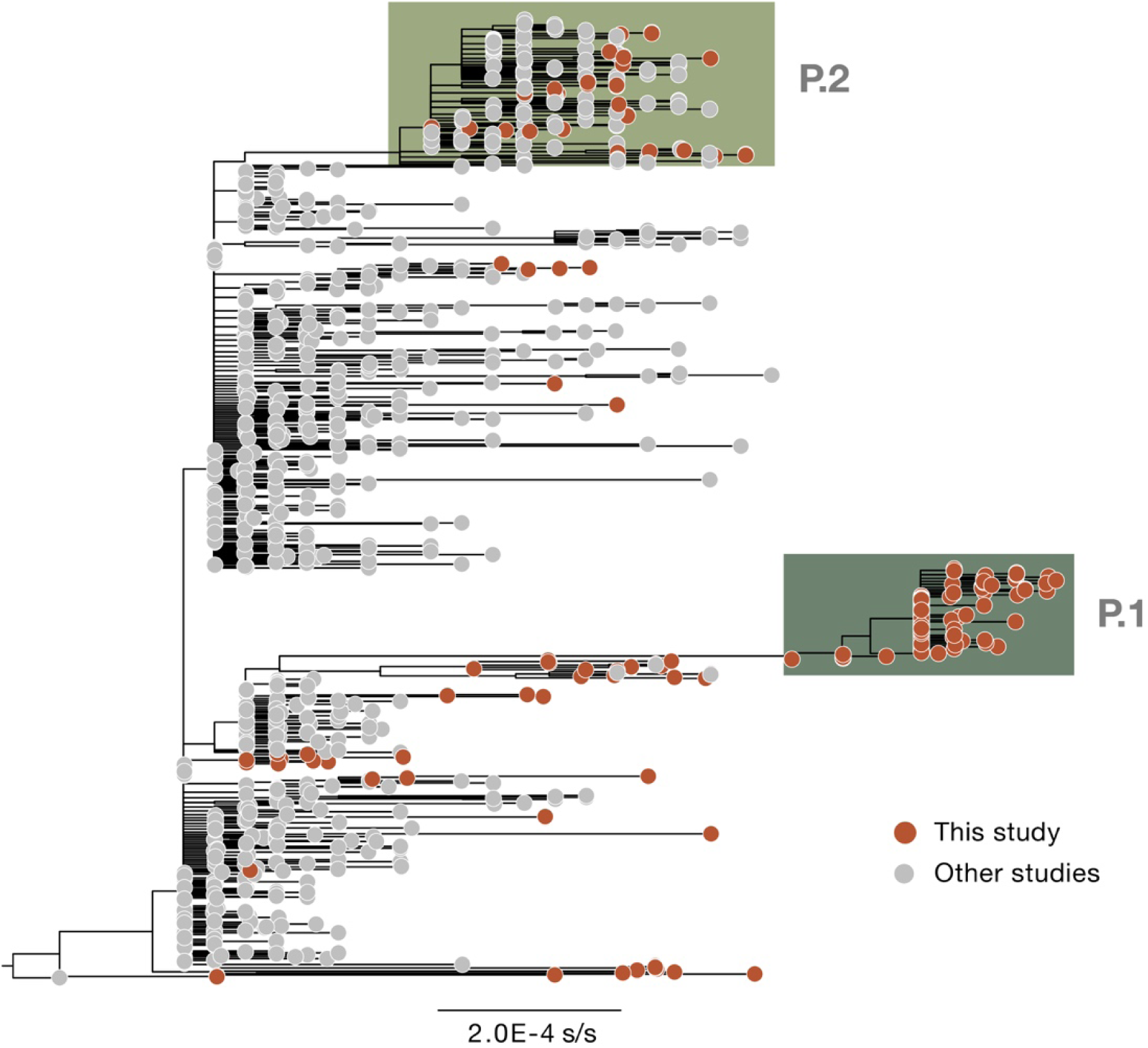
Maximum likelihood tree estimated for *dataset A’* (*n*=999). This dataset was used to confirm lineage assignment for all sequences generated in this study regardless of genome coverage (see also fig. S1). s/s=nucleotide substitutions per site.

**Figure S3.**
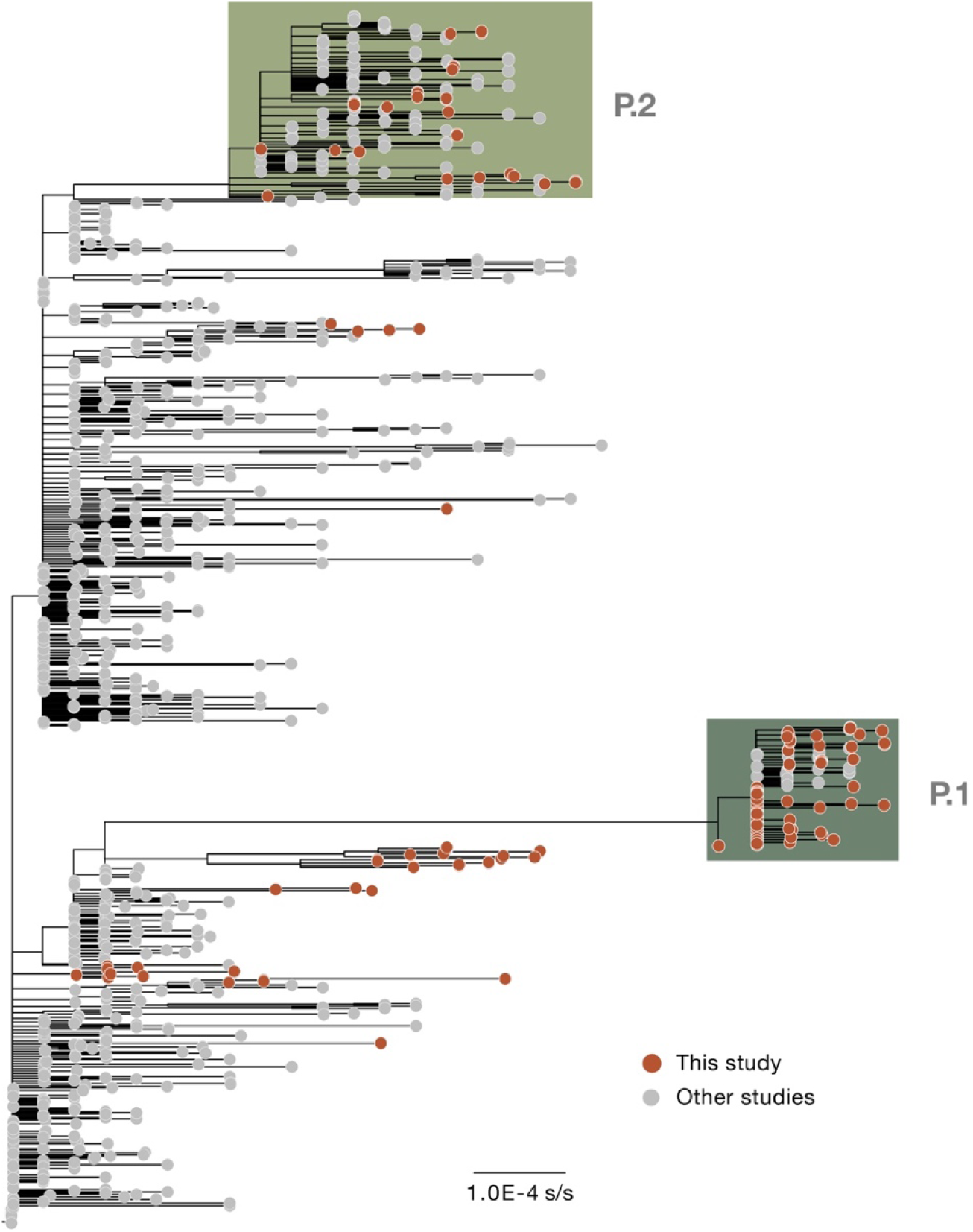
Maximum likelihood tree estimated for *dataset B’* (*n*=974). s/s=nucleotide substitutions per site. This phylogeny includes only publicly available and published sequences classified as B.1.1.28, P.1 and P.2 lineages.

**Figure S4.**
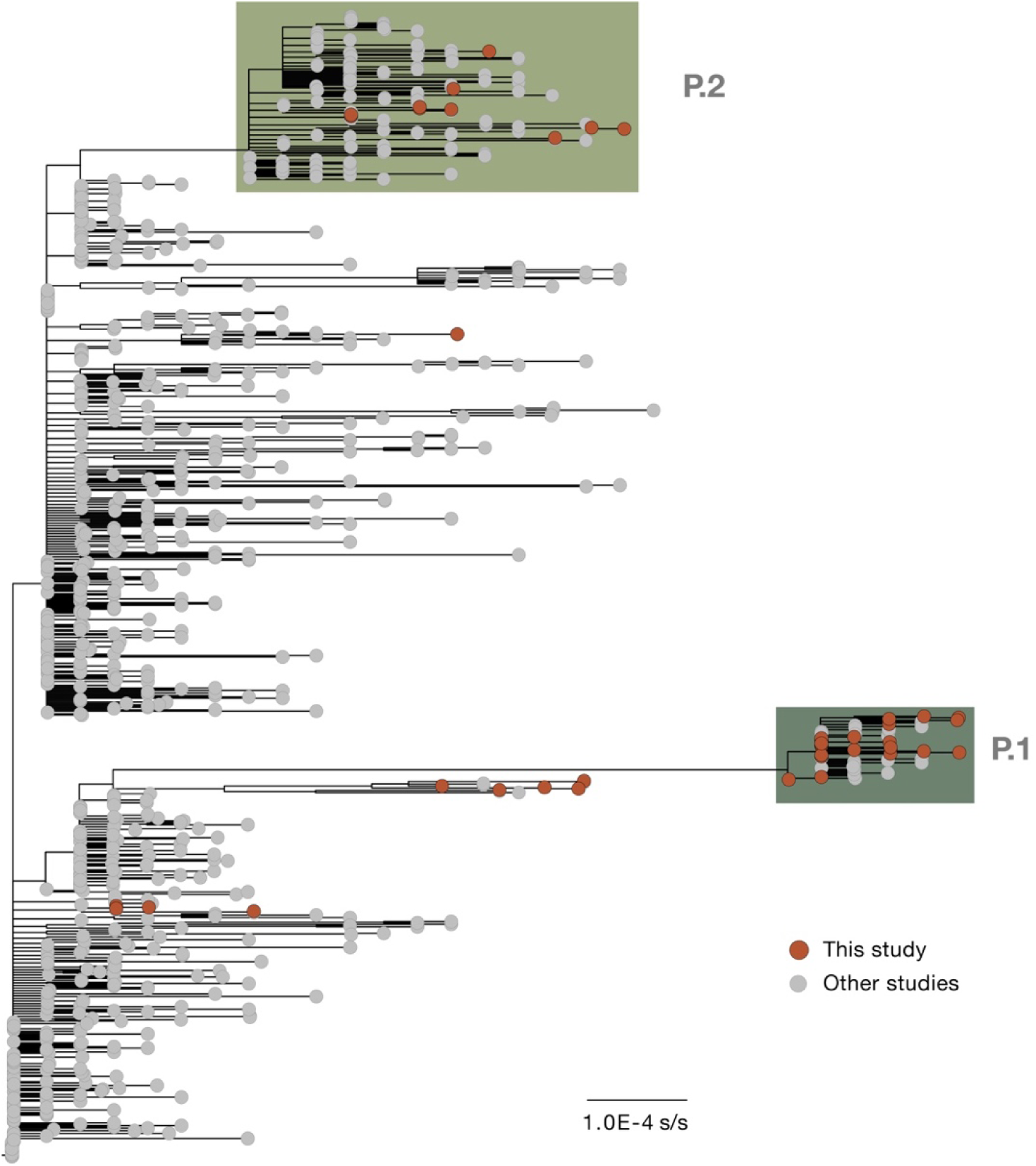
Maximum likelihood tree estimated for *dataset C’* (*n*=883). s/s=nucleotide substitutions per site. This phylogeny includes only publicly available and published sequences classified as B.1.1.28, P.1 and P.2 lineages.

**Figure S5.**
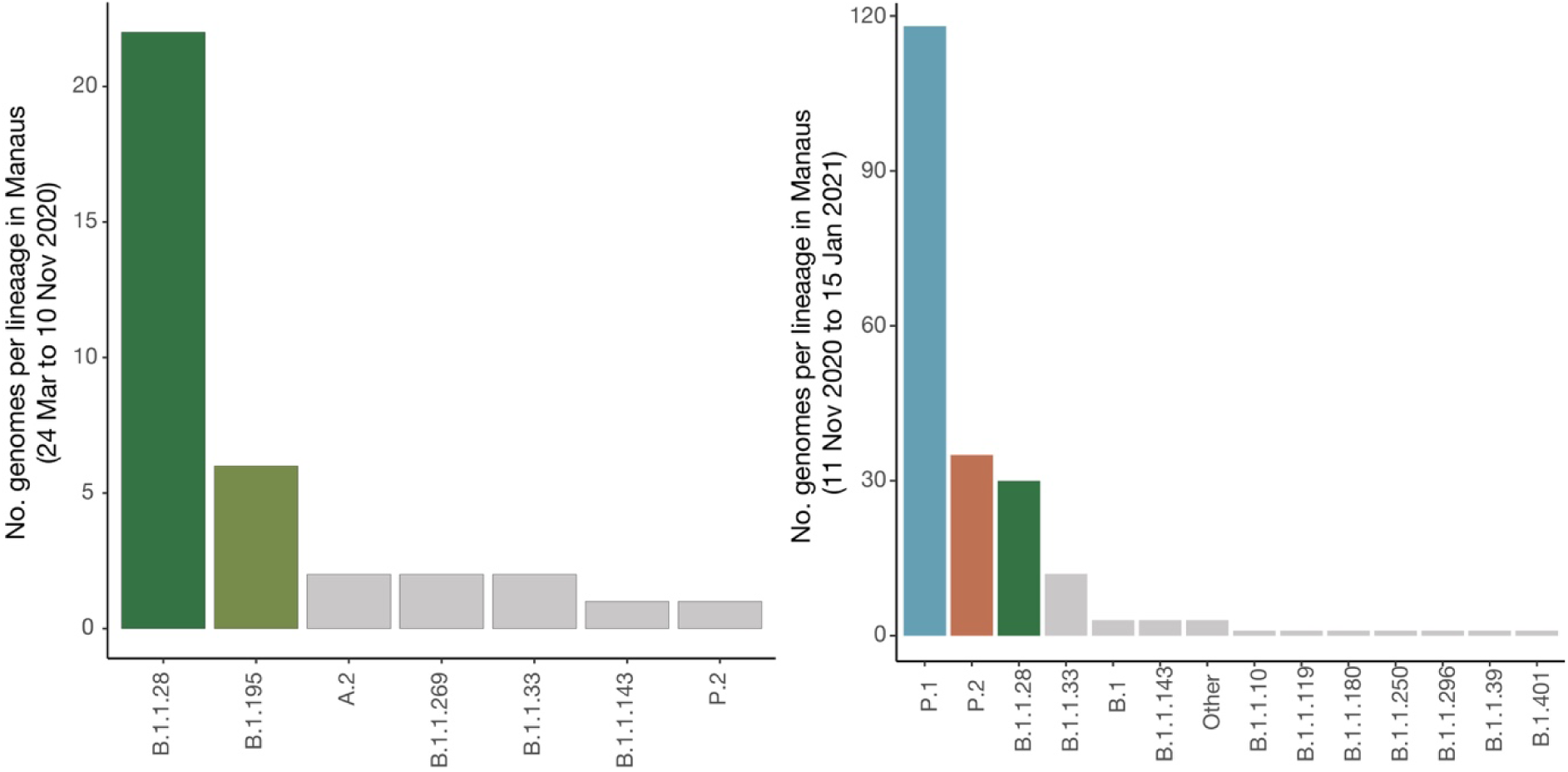
Pango lineages identified in Manaus among our 184 sequences samples and publicly available genomes in GISAID between 24 March and 10 November 2020 (left panel) and between 11 November and 15 January 2020 (right panel). Duplicate sequences and sequences with no date or location of sample collection were removed. Coloured bars correspond to lineages that represent >10% of sequenced samples during each time period.

**Fig. S6.**
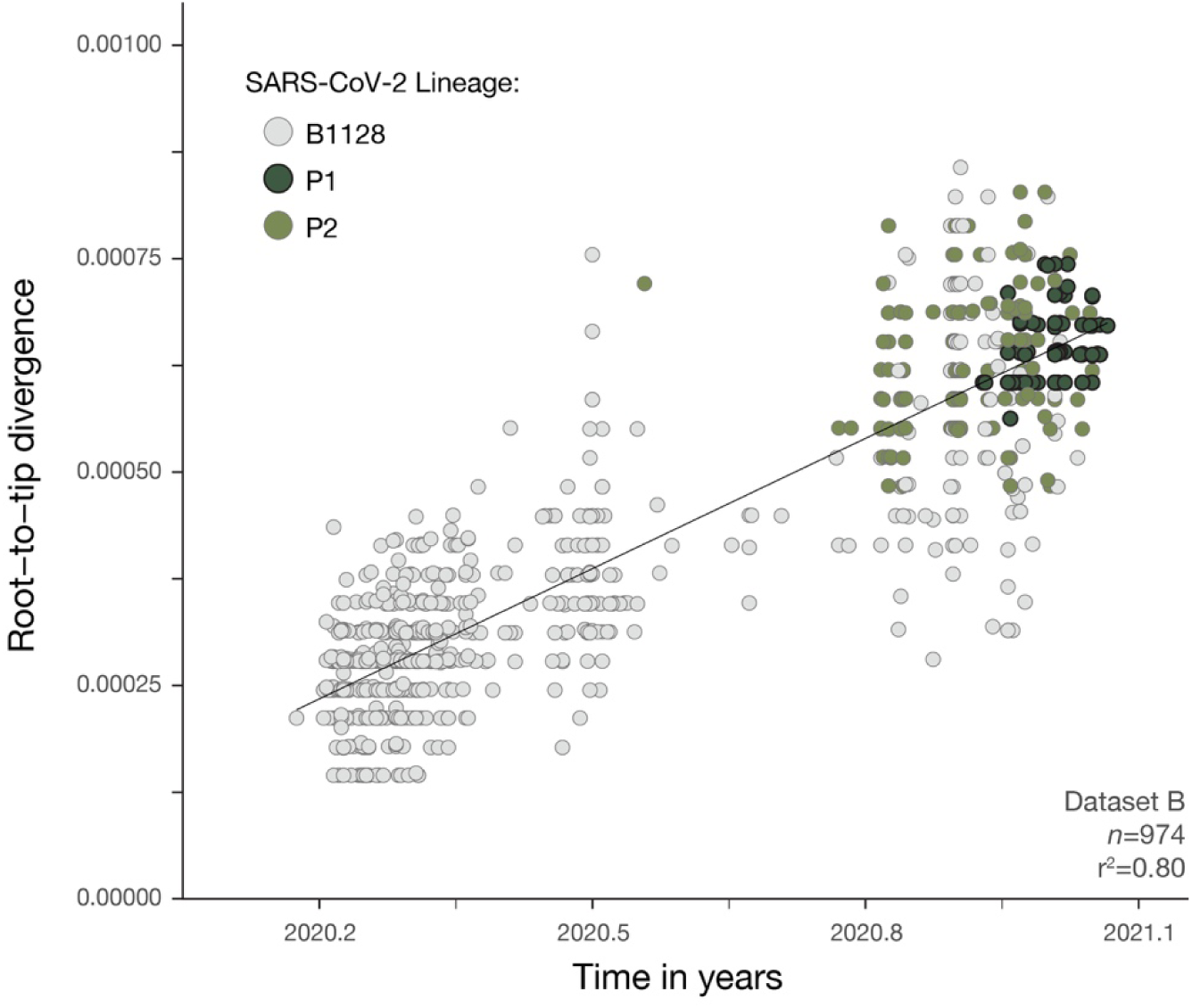
Regression of root-to-tip genetic distances and sampling dates for *dataset B’* estimated using TempEst v.1.5.3 (*27*). Circles corresponds to the tips of the maximum likelihood phylogenetic tree show in fig. S3.

**Fig. S7.**
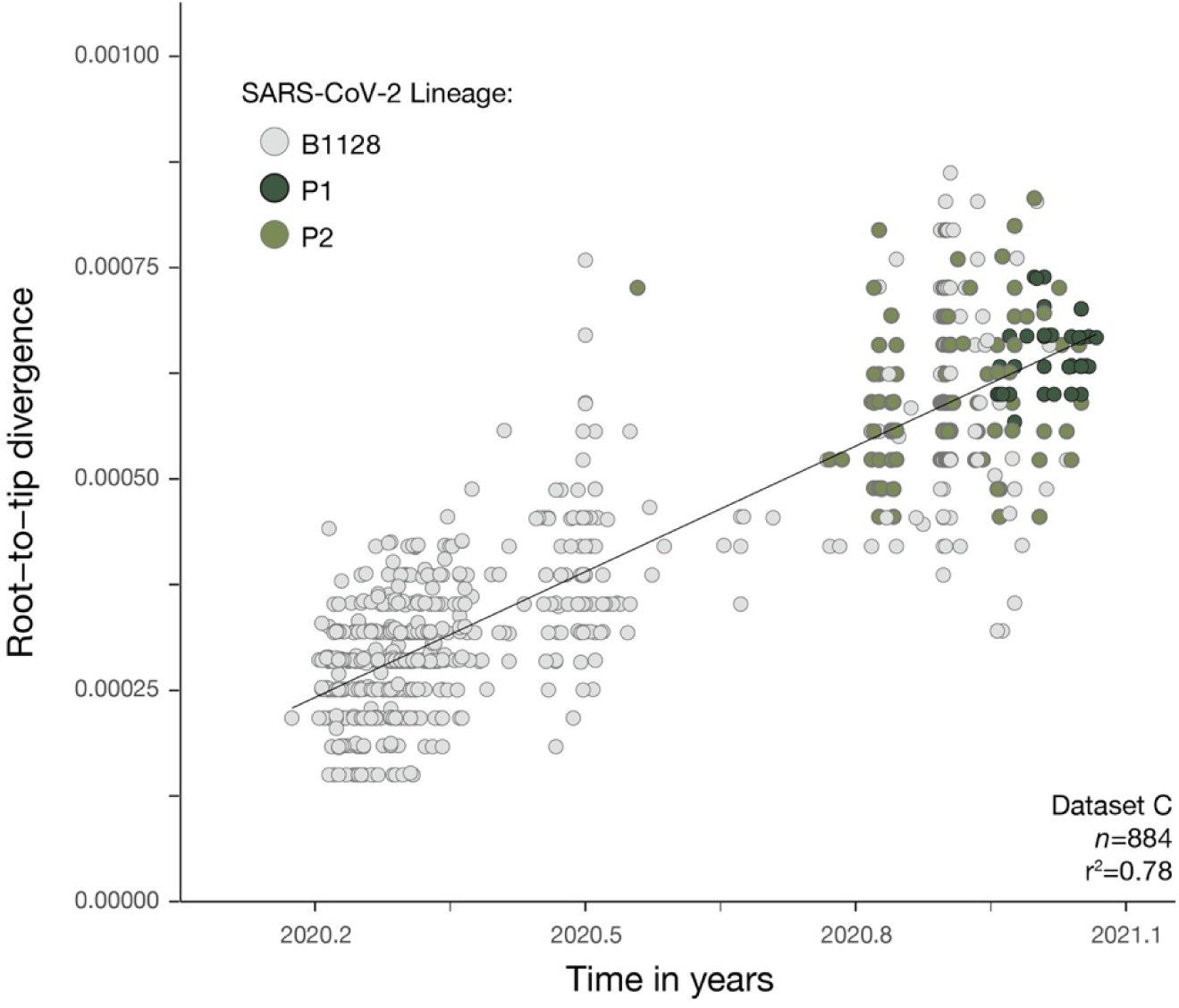
Regression of root-to-tip genetic distances and sampling dates for *dataset C’* estimated using TempEst v.1.5.3 (*27*). Circles corresponds to the tips of the maximum likelihood phylogenetic tree show in fig. S4.

**Fig. S8.**
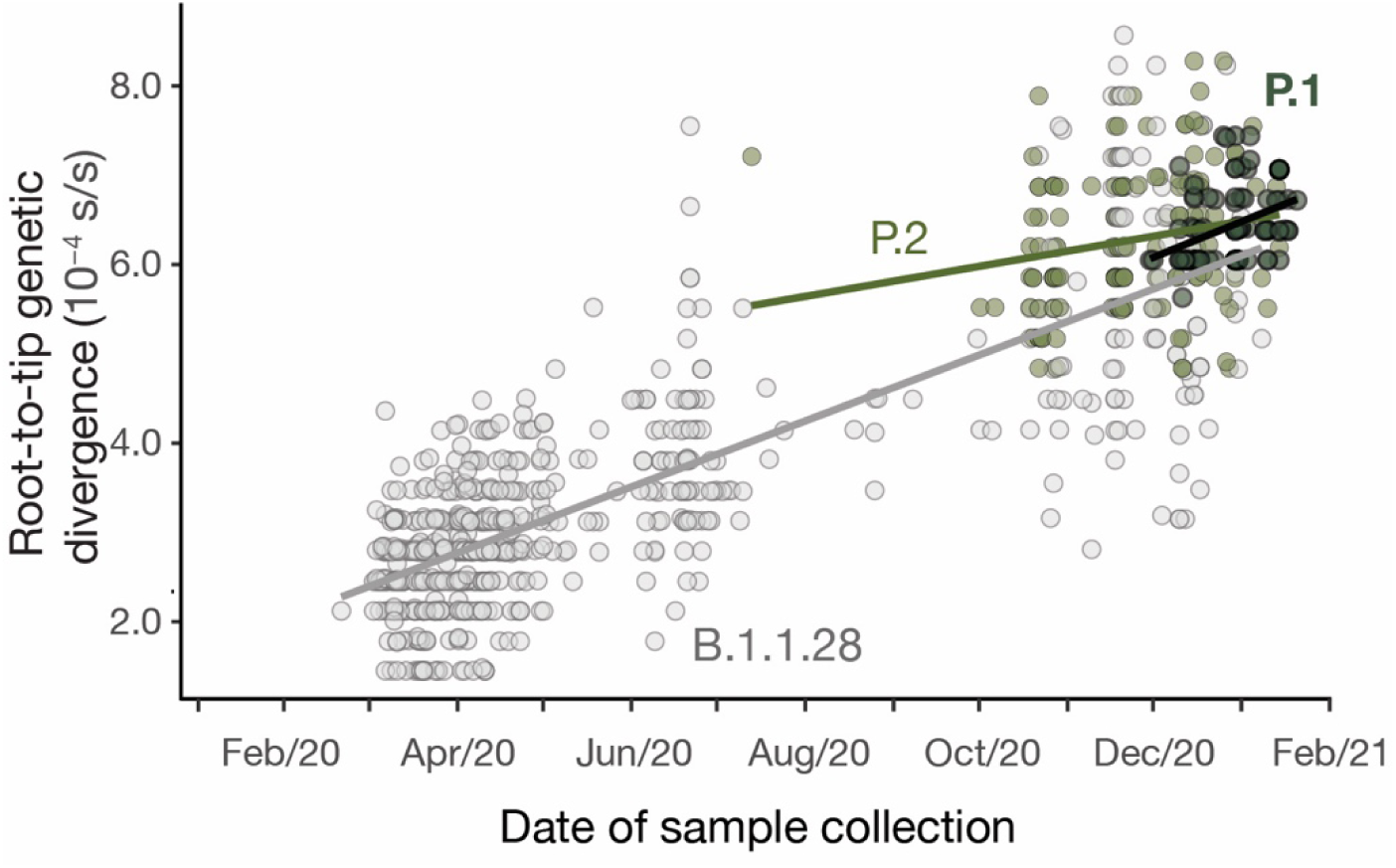
Regression of root-to-tip genetic distances and sampling dates for *dataset B’* estimated using TempEst v.1.5.3 (*27*), with separate regression lines for B.1.1.28 and P.1 lineages computed in R v 3.6.2 (*54*). Circles corresponds to the tips of the maximum likelihood phylogenetic tree show in fig. S3.

**Fig. S9.**
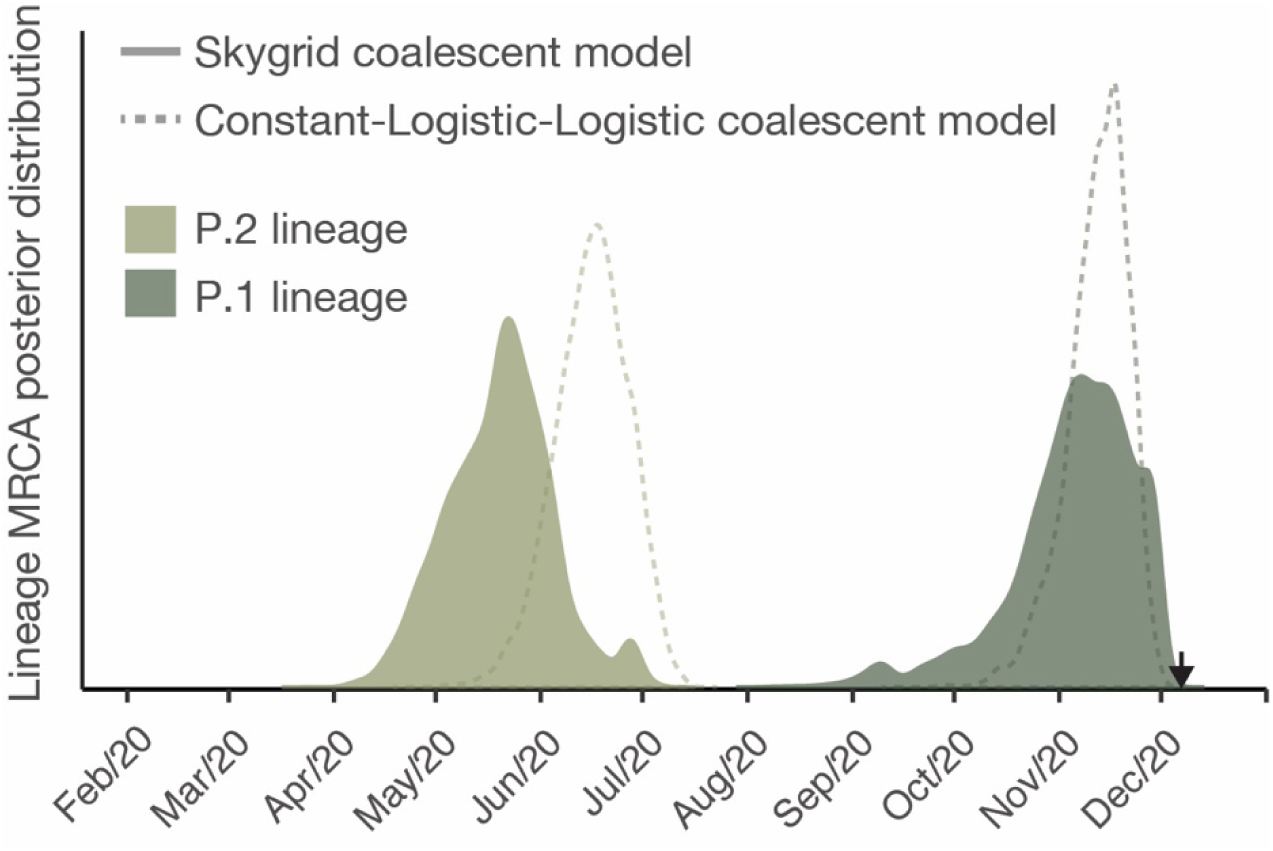
Posterior estimates of the time of the most recent common ancestor of P.1 (dark green) and P.2 (light green) lineages estimated using a flexible non-parametric skygrid coalescent model (*31*). Dashed lines show the posterior estimates for the same evolutionary parameters but estimated using a constant-logistic-logistic model (see Materials and Methods for details). Both coalescent models are implemented in BEAST v.1.10 (*55*).

**Fig. S10.**
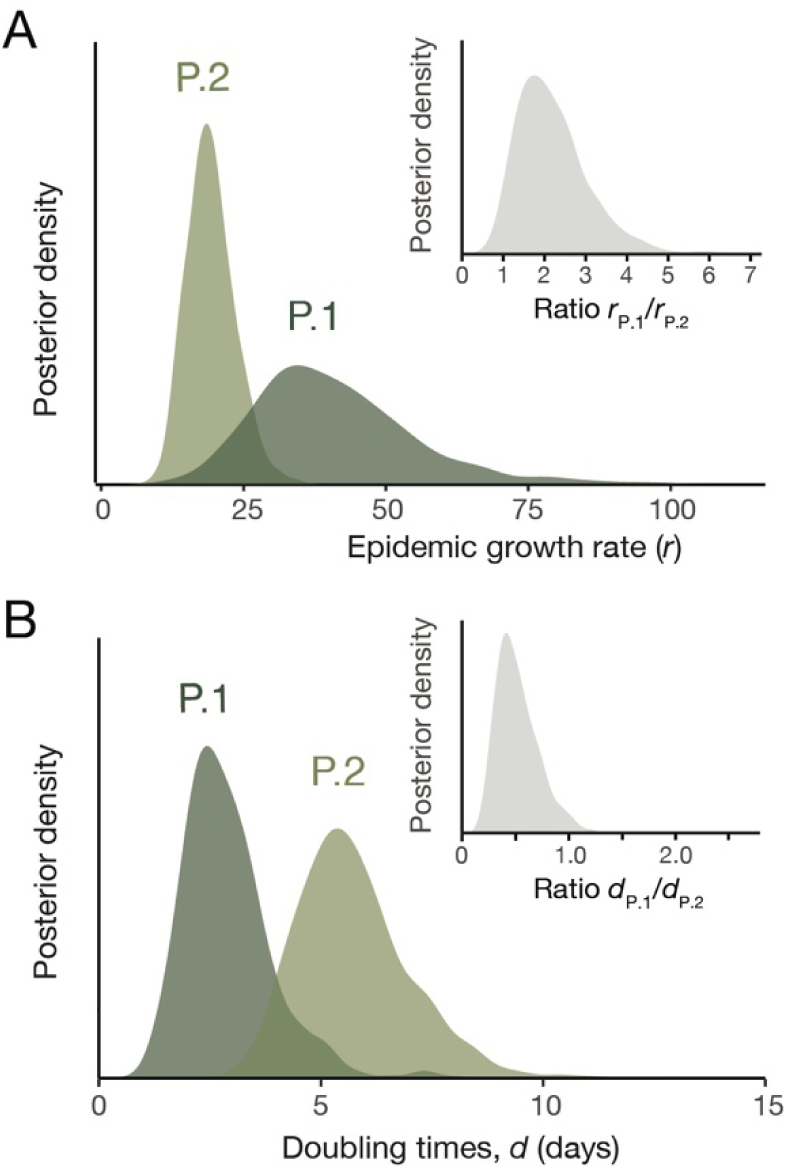
Coalescent growth rates for P.1 and P.2 lineages estimated using a constant-logistic-logistic approach implemented in BEAST v.1.10 (*55*). (**A**) Light and dark green posterior probability distributions show virus lineage population growth (*r*) for P.2 and P.1, respectively. Inset shows posterior probability estimates for the ratio of epidemic growth rates between P.1 and P.2 (**B**) Light and dark green posterior probability distributions of the estimated doubling times for P.2 and P.1 lineage, respectively. Inset shows posterior probability estimates for the ratio of doubling times between P.1 and P.2.

**Fig. S11.**
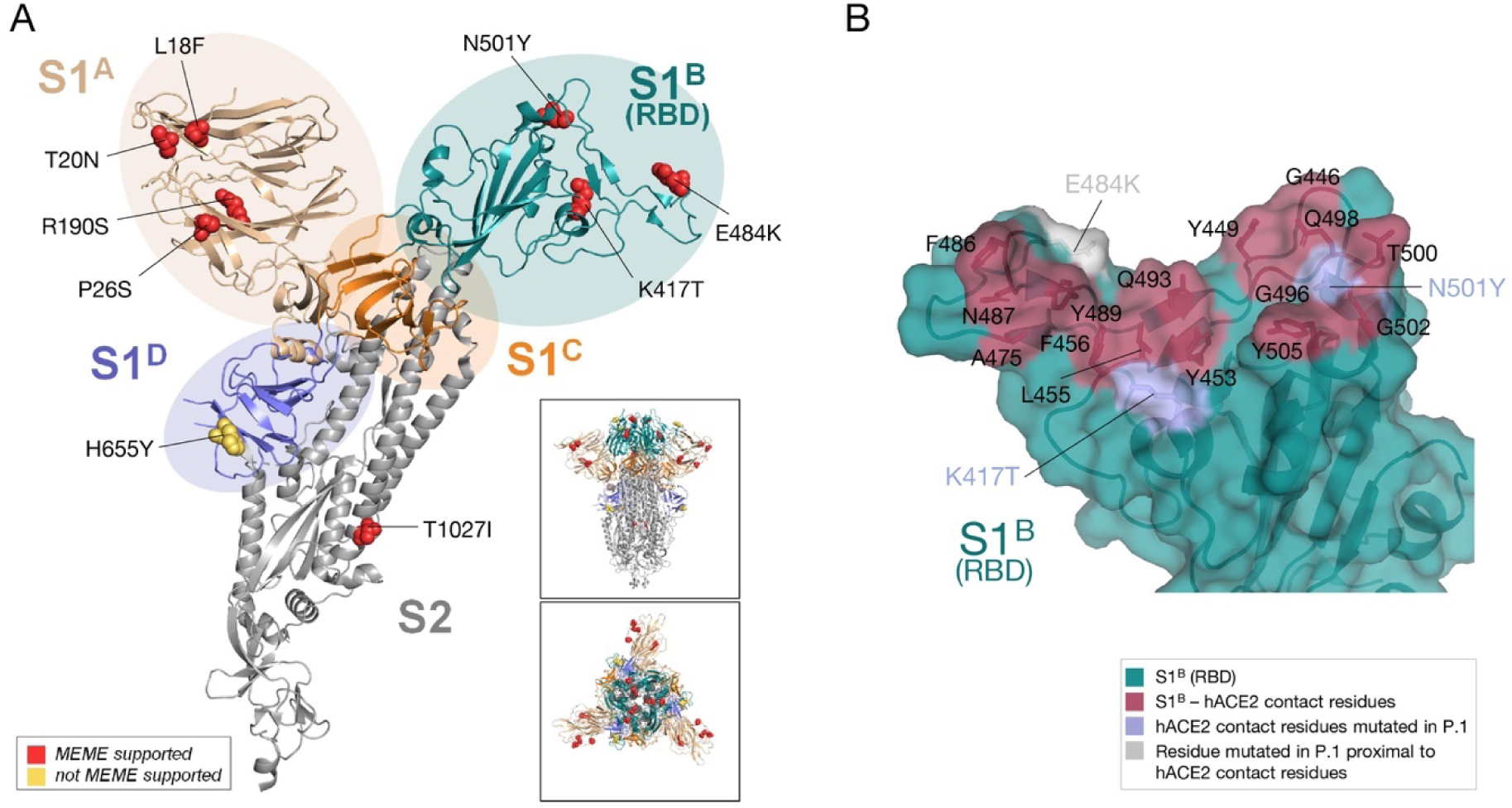
Mapping of adaptive substitutions onto the structure of SARS-COV-2 S. **(A)** Lineage defining mutations within the S protein of the P.1 lineage are mapped onto the spike glycoprotein structure of SARS-CoV-2 (*62*) (*NSMB;* PDB: 6ZGI). Cartoon representation of one protomer of the trimeric S ectodomain structure with the different domains and subunits indicated by color: S1^A^ (wheat), S1^B^ (RBD, teal), S1^C^ (orange), S1^D^ (blue), S2 subunit (grey). Residues under selection are shown as spheres with associated mutations indicated and colored according to the respective type of selection as analyzed under MEME (*60*) and supported for at least one branch (red) or not supported (yellow). The positively selected residue V1176F was not resolved in the cryoEM map of the SARS-CoV-2 S ectodomain used here. The inset panels reflect the same presentation in the trimeric context of the S protein in a side (*upper panel*) and top-down view (*lower panel*). N-linked glycans are omitted for clarity. (**B)** Surface representation of the SARS-CoV-2 S RBD-hACE2 contact interface with residues mutated in the RBD of P.1 highlighted. Contact residues as observed in the SARS-CoV-2 S RBD (deep teal) in complex with hACE2 ((*11*) PDB: 6MOJ) are colored red with side chains shown as sticks. Residues mutated in lineage P.1 that are part of the contact interface (K417 and N501) are colored light blue. A nearby residue that is observed to be mutated in P.1 that is not part of the direct contact interface (E484) is colored grey with side chains shown as sticks.

**Fig. S12.**
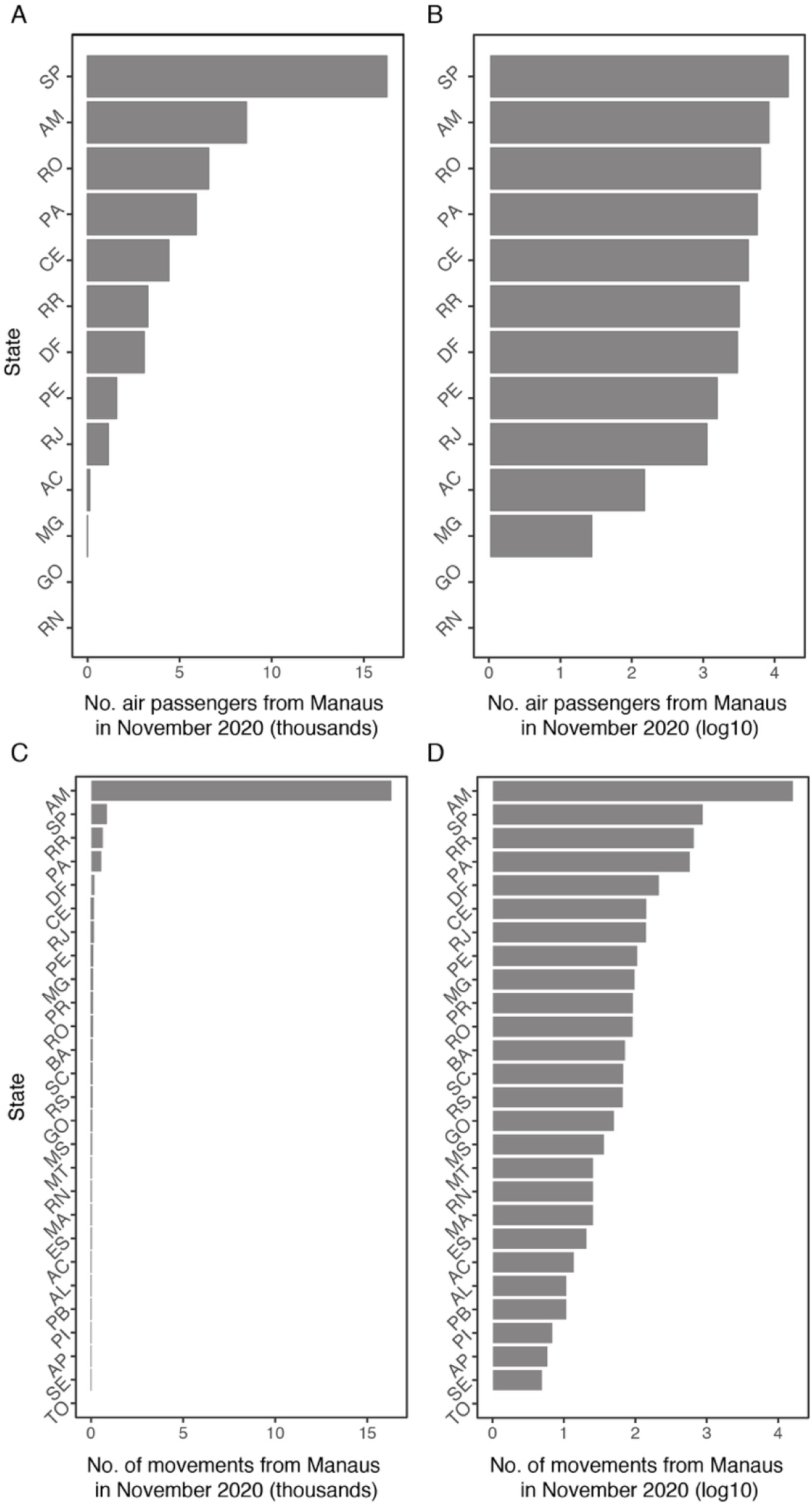
Number of movements from Manaus to federal units in Brazil obtained from ANAC flight data (A and B) and from anonymized cell phone data (C and D). X-axis of panels B and D are shown in log_10_ units. The ISO 3166-2:BR codes of the states AC–Acre, AL–Alagoas, AP– Amapá, AM–Amazonas, BA–Bahia, CE–Ceará, DF–Distrito Federal, ES–Espírito Santo, GO–Goiás, MA–Maranhão, MT–Mato Grosso, MS–Mato Grosso do Sul, MG–Minas Gerais, PA–Pará, PB–Paraíba, PR–Paraná, PE–Pernambuco, PI–Piauí, RJ–Rio de Janeiro, RN–Rio Grande do Norte, RS–Rio Grande do Sul, RO–Rondônia, RR–Roraima, SC–Santa Catarina, SP–São Paulo, SE–Sergipe, TO–Tocantins.

**Table S1.** Proportion of P.1 cases in Manaus. Note that week commencing on 30 Nov 2020 includes the date of the first P.1 case detected in our study (6 Dec 2020).

| **Date (Week Commencing)** | **No. Sequenced** | **No. P.1** | **Proportion** |
| --- | --- | --- | --- |
| 2 November 2020 | 9 | 0 | 0 |
| 9 November 2020 | 2 | 0 | 0 |
| 16 November 2020 | 4 | 0 | 0 |
| 23 November 2020 | NA | NA | NA |
| 30 November 2020 | 2 | 1 | 50% |
| 7 December 2020 | 7 | 1 | 14% |
| 14 December 2020 | 24 | 7 | 29% |
| 21 December 2020 | 37 | 20 | 54% |
| 28 December 2020 | 14 | 8 | 57% |
| 4 January 2021 | 46 | 40 | 87% |

**Table S2.** Positively selected sites in P.1 lineage defined with statistical support (MEME) for at least one branch.

| Nucleotide | Gene/Site | Signature | dN/dS>1 (MEME) | P.1 substitution to |
| --- | --- | --- | --- | --- |
| 1216 | ORF1A/318 |  |  | L |
| 3115 | ORF1A/951 |  | Yes | I |
| 3265 | ORF1A/1001 |  |  |  |
| 3826 | ORF1A/1188 | Yes | Yes | L |
| 5647 | ORF1A/1795 | Yes | Yes | Q |
| 6952 | ORF1A/2230 |  |  |  |
| 7069 | ORF1A/2269 |  |  |  |
| 10666 | ORF1A3468 |  |  |  |
| 11422 | ORF1A/3720 |  | Yes | N |
| 12052 | ORF1A/3930 |  |  |  |
| 14119 | ORF1B/218 |  | Yes | L |
| 21613 | S/18 | Yes | Yes | F |
| 21619 | S/20 | Yes | Yes | N |
| 21637 | S/26 | Yes | Yes | S |
| 22129 | S/190 | Yes | Yes | S |
| 22810 | S/417 | Yes | Yes | T |
| 23011 | S/484 | Yes |  | K |
| 23062 | S/501 | Yes | Yes | Y |
| 23437 | S/626 |  |  |  |
| 23524 | S/655 | Yes |  |  |
| 23623 | S/688 |  |  |  |
| 24640 | S/1027 | Yes | Yes | I |
| 25087 | S/1176 | Yes | Yes | F |
| 25521 | ORF3A/44 |  |  |  |
| 26148 | ORF3A/253 | Yes | Yes | P |
| 28510 | N/80 | Yes | Yes | R |
| 28627 | N/119 |  |  |  |
| 28876 | N/202 |  | Yes | S |
| 28984 | N/238 |  |  |  |
| 29194 | N/308 |  |  |  |

**Table S3.**
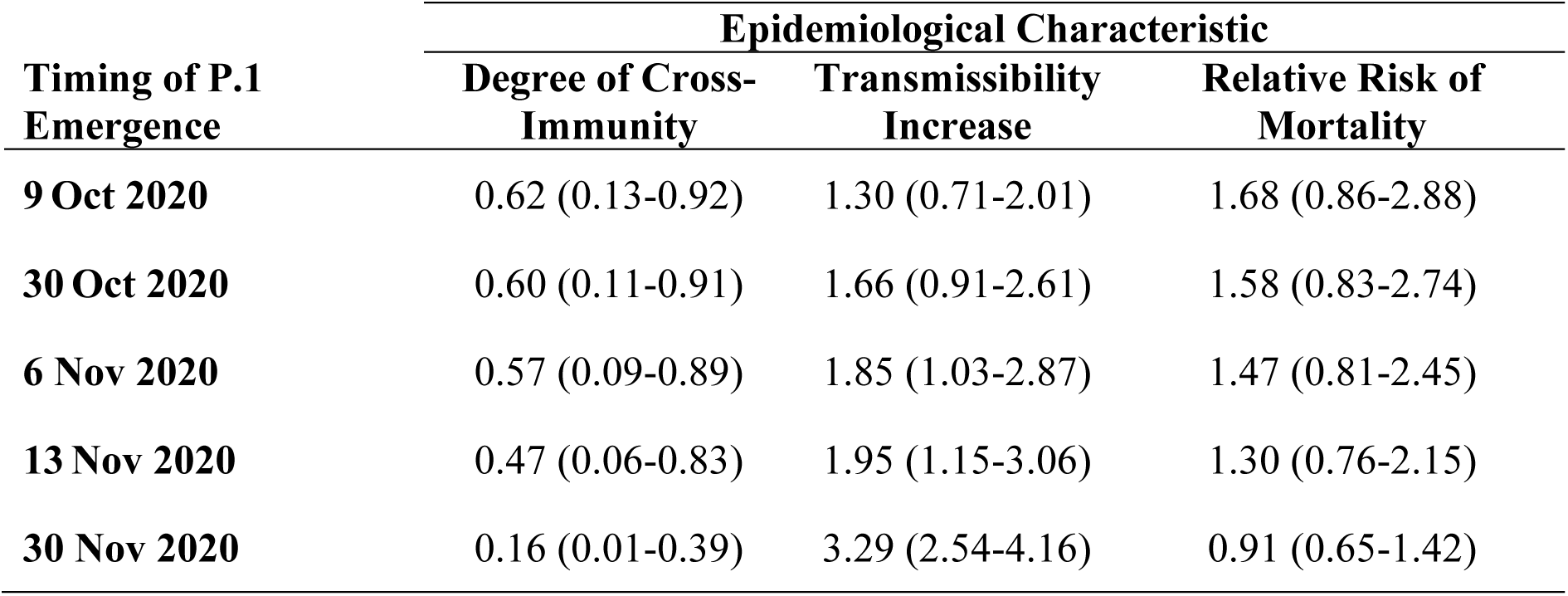
Inferred changes in epidemiological characteristics of P.1, depending on the timing of P.1 emergence assumed. Results presented are the mean, with the 95% Bayesian Credible Interval, BCI, in brackets). Note that “Degree of Cross-Immunity” is equal to 1 – the “Immune Evasion” described in the main text.

| **Timing of P.1**<br>**Emergence** | **Epidemiological Characteristic** |  |  |
| --- | --- | --- | --- |
|  | **Degree of Cross-**<br>**Immunity** | **Transmissibility**<br>**Increase** | **Relative Risk of**<br>**Mortality** |
| **9 Oct 2020** | 0.62 (0.13-0.92) | 1.30 (0.71-2.01) | 1.68 (0.86-2.88) |
| **30 Oct 2020** | 0.60 (0.11-0.91) | 1.66 (0.91-2.61) | 1.58 (0.83-2.74) |
| **6 Nov 2020** | 0.57 (0.09-0.89) | 1.85 (1.03-2.87) | 1.47 (0.81-2.45) |
| **13 Nov 2020** | 0.47 (0.06-0.83) | 1.95 (1.15-3.06) | 1.30 (0.76-2.15) |
| **30 Nov 2020** | 0.16 (0.01-0.39) | 3.29 (2.54-4.16) | 0.91 (0.65-1.42) |

**Table S4.** Inferred changes in epidemiological characteristics of P.1, depending on the rate of natural immunity waning assumed (for an emergence date of 6 Nov 2020). Results presented are the mean, with the 95% Bayesian Credible Interval in brackets). Note that “Degree of Cross-Immunity” is equal to 1 – the “Immune Evasion” described in the main text.

| **Duration of Immunity Assumed** | **Epidemiological Characteristic** |  |  |
| --- | --- | --- | --- |
|  | **Degree of Cross-Immunity** | **Transmissibility Increase** | **Relative Risk of Mortality** |
| 50% waning over 8-month period | 0.42 (0.04-0.81) | 1.58 (0.98-2.51) | 1.37 (0.89-2.10) |
| 50% waning over 1-year period | 0.57 (0.09-0.89) | 1.85 (1.03-2.87) | 1.47 (0.81-2.45) |
| 50% waning over 18-month period | 0.62 (0.15-0.91) | 1.96 (1.14-2.97) | 1.50 (0.74-2.65) |

**Data S1. (separate file)**

Metadata and GISAID IDs for 1,084 SARS-CoV-2 samples from Manaus.

**Data S2. (separate file)**

Metadata and GISAID IDs for 184 sequenced samples from Manaus.

**Data S3. (separate file)**

GISAID Acknowledgment table (used for datasets B and C).

**Data S4. (separate file)**

Flight mobility data from Manaus (state level).

**Data S5. (separate file)**

Cell-phone derived mobility data from Manaus (municipality level).

## References

1. P. C. Hallal, et al., SARS-CoV-2 antibody prevalence in Brazil: results from two successive nationwide serological household surveys. The Lancet Global Health 8, e1390–e1398 (2020).

2. L. F. Buss et al., Three-quarters attack rate of SARS-CoV-2 in the Brazilian Amazon during a largely unmitigated epidemic. Science 371, 288–292 (2021).

3. M.-S. G. Álvarez-Antonio C, Calampa C, Casanova W, Carey C, Alava F, Rodríguez-Ferrucci H, Quispe AM, Seroprevalence of Anti-SARS-CoV-2 Antibodies in Iquitos, Loreto, Peru. medRxiv, (2021).

4. M. Mercado, Ospina, M., Institutoo Nacional de Salud, “Seroprevalencia de SARS-CoV-2 durante la epidemia en Colombia: estudio país” (https://www.ins.gov.co/BibliotecaDigital/Seroprevalencia-estudio-colombia.pdf, 2020).

5. Portal da Fundação de Vigilância em Saúde do Amazonas, “Perfil clínico e demográfico dos casos de Covid-19 no estado do Amazonas: uma análise comparativa entre 2020 e 2021,” No. 17 (2021)

6. A. J. Greaney, Loes, A. N., Crawford, K. H. D., Starr, T. N., Malone, K. D., Chu, H. Y., Bloom, J. D., Comprehensive mapping of mutations to the SARS-CoV-2 receptor-binding domain that affect recognition by polyclonal human serum antibodies. Cold Spring Harbor Laboratory p. 2020.12.31.425021 (2021).

7. T. N. Starr et al., Deep Mutational Scanning of SARS-CoV-2 Receptor Binding Domain Reveals Constraints on Folding and ACE2 Binding. Cell 182, 1295–1310 e1220 (2020).

8. M. A. Suchard, R. E. Weiss, J. S. Sinsheimer, Testing a molecular clock without an outgroup: derivations of induced priors on branch-length restrictions in a Bayesian framework. Systematic Biology 52, 48–54 (2003).

9. Z. Wang et al., mRNA vaccine-elicited antibodies to SARS-CoV-2 and circulating variants. Nature (2021).

10. Y. Weisblum et al., Escape from neutralizing antibodies by SARS-CoV-2 spike protein variants. eLife 9 (2020).

11. J. Lan et al., Structure of the SARS-CoV-2 spike receptor-binding domain bound to the ACE2 receptor. Nature 581, 215–220 (2020).

12. E. W. H. Tegally, M. Giovanetti, A. Iranzadeh, V. Fonseca, J. Giandhari, D. Doolabh, S. Pillay, E. J. San, N. Msomi, K. Mlisana, A. von Gottberg, S. Walaza, M. Allam, A. Ismail, T. Mohale, A. J. Glass, S. Engelbrecht, G. Van Zyl, W. Preiser, F. Petruccione, Sigal D. Hardie, G. Marais, M. Hsiao, S. Korsman, M.-A. Davies, L. Tyers, I. Mudau, D. York, C. Maslo, D. Goedhals, S. Abrahams, O. Laguda-Akingba, A. Alisoltani-Dehkordi, A. Godzik, C. K. Wibmer, B. T. Sewell, J. Lourenço, L. C. J. Alcantara, S. L. K. Pond, S. Weaver, D. Martin, R. J. Lessells, J. N. Bhiman, C. Williamson, T. de Oliveira, Emergence and rapid spread of a new severe acute respiratory syndrome-related coronavirus 2 (SARS-CoV-2) lineage with multiple spike mutations in South Africa. bioRxiv (https://doi.org/10.1101/2020.12.21.20248640, 2021).

13. A. Rambaut, Loman, N., Pybus, O., Barclay, W., Barrett, J., Carabelli., A., Connor, T., Peacock, T., Robertson, D. L., Volz, E., on behalf of COVID-19 Genomics Consortium UK (CCoG-UK), “Preliminary genomic characterisation of an emergent SARS-CoV-2 lineage in the UK defined by a novel set of spike mutations” (https://virological.org/t/preliminary-genomic-characterisation-of-an-emergent-sars-cov-2-lineage-in-the-uk-defined-by-a-novel-set-of-spike-mutations/563, 2020).

14. S. A. M. R. Council, Report on Weekly Deaths in South Africa (available at https://www.samrc.ac.za/reports/report-weekly-deaths-south-africa?bc=254). (2021).

15. N. L. Washington et al., Genomic epidemiology identifies emergence and rapid transmission of SARS-CoV-2 B.1.1.7 in the United States. medRxiv (https://doi.org/10.1101/2021.02.06.21251159, 2021).

16. D. D. S. Candido et al., Routes for COVID-19 importation in Brazil. J Travel Med 27, (2020).

17. D. S. Candido et al., Evolution and epidemic spread of SARS-CoV-2 in Brazil. Science 369, 1255–1260 (2020).

18. W. M. de Souza et al., Epidemiological and clinical characteristics of the COVID-19 epidemic in Brazil. Nat Hum Behav 4, 856–865 (2020).

19. J. G. Jesus et al., Importation and early local transmission of COVID-19 in Brazil, 2020. Revista do Instituto de Medicina Tropical de Sao Paulo 62, e30 (2020).

20. I. M. Claro et al., Local Transmission of SARS-CoV-2 Lineage B.1.1.7, Brazil, December 2020. Emerging infectious diseases 27, (2021).

21. V. A. D. Nascimento et al., Genomic and phylogenetic characterisation of an imported case of SARS-CoV-2 in Amazonas State, Brazil. Memorias do Instituto Oswaldo Cruz 115, e200310 (2020).

22. J. R. Tyson et al., Improvements to the ARTIC multiplex PCR method for SARS-CoV-2 genome sequencing using nanopore. bioRxiv (https://doi.org/10.1101/2020.09.04.283077, 2020).

23. WHO, “Genomic sequencing of SARS-CoV-2 - A guide to implementation for maximum impact on public health” (https://www.who.int/publications/i/item/9789240018440, Geneva, 2021).

24. A. Rambaut et al., A dynamic nomenclature proposal for SARS-CoV-2 lineages to assist genomic epidemiology. Nat Microbiol 5, 1403–1407 (2020).

25. N. R. Faria, Claro, I. M., Candido, D., Franco, L. A. M., P. S. Andrade, T. M. Coletti, C. M. Silva, F. C. Sales, E. R. Manuli, R. S. Aguiar, N. Gaburo, C. C. Camilo, N. A. Fraiji, M. A. E. Crispim, M. P. S. S. Carvalho, A. Rambaut, N. Loman, O. G. Pybus, E. C. Sabino, on behalf of CADDE Genomic Network, “Genomic characterisation of an emergent SARS-CoV-2 lineage in Manaus: preliminary findings” (https://virological.org/t/genomic-characterisation-of-an-emergent-sars-cov-2-lineage-in-manaus-preliminary-findings/586, 2021).

26. T. Fujino et al., Novel SARS-CoV-2 Variant Identified in Travelers from Brazil to Japan. Emerging Infectious Diseases 27, (2021).

27. A. Rambaut, T. T. Lam, L. Max Carvalho, O. G. Pybus, Exploring the temporal structure of heterochronous sequences using TempEst (formerly Path-O-Gen). Virus Evol 2, vew007 (2016).

28. B. Choi et al., Persistence and Evolution of SARS-CoV-2 in an Immunocompromised Host. The New England Journal of Medicine 383, 2291–2293 (2020).

29. V. A. Avanzato et al., Case Study: Prolonged Infectious SARS-CoV-2 Shedding from an Asymptomatic Immunocompromised Individual with Cancer. Cell 183, 1901–1912 e1909 (2020).

30. A. J. Drummond, M. A. Suchard, Bayesian random local clocks, or one rate to rule them all. BMC Biology 8, 114 (2010).

31. M. S. Gill et al., Improving Bayesian population dynamics inference: a coalescent-based model for multiple loci. Molecular Biology and Evolution 30, 713–724 (2013).

32. M. Marks et al., Transmission of COVID-19 in 282 clusters in Catalonia, Spain: a cohort study. The Lancet Infectious Diseases (2021).

33. J. A. Hay, Kennedy-Shaffer, L., Kanjilal, S., Lipsitch, M., Mina, M. J, Estimating epidemiologic dynamics from single cross-sectional viral load distributions. medRxiv 2021 (https://doi.org/10.1101/2020.10.08.20204222, 2021).

34. A. R. M. Kidd, A. Best, J. Mirza, B. Percival, M. Mayhew, O. Megram, F. Ashford, T. White, E. Moles-Garcia, L. Crawford, A. Bosworth, T. Plant, A. McNally, S-variant SARS-CoV-2 is associated with significantly higher viral loads in samples tested by ThermoFisher TaqPath RT-QPCR. bioRxiv (https://doi.org/10.1101/2020.12.24.20248834, 2020).

35. K. Stephen, Fauver, J. R., Mack, C., Tai, C. G., Breban, M. I, et al., Densely sampled viral trajectories suggest longer duration of acute infection with B.1.1.7 variant relative to non-B.1.1.7 SARS-CoV-2. https://nrs.harvard.edu/URN-3:HUL.INSTREPOS:37366884 (2021).

36. E. C. Sabino et al., Resurgence of COVID-19 in Manaus, Brazil, despite high seroprevalence. Lancet 397, 452–455 (2021).

37. V. Hall, Foulkes, S., Charlett, A., Atti, A., Monk, E. J. M., Simmons, R., Wellington, E., et al., Do antibody positive healthcare workers have lower SARS-CoV-2 infection rates than antibody negative healthcare workers? Large multi-centre prospective cohort study (the SIREN study), England: June to November 2020. bioRxiv, (2021).

38. S. F. McGough, M. A. Johansson, M. Lipsitch, N. A. Menzies, Nowcasting by Bayesian Smoothing: A flexible, generalizable model for real-time epidemic tracking. PLoS Computational Biology 16, e1007735 (2020).

39. I. Hawryluk, Hoeltgebaum, H., Mishra, S., Miscouriduo, X., Schnekenberg, R. P., Whittaker, C., Vollmer, M., Flaxman, S., Mellaan, T. A., Gaussian Process Nowcasting: Application to COVID-19 Mortality Reporting. arXiv arXiv:2102.11249, (2021).

40. Agência Brasil. Covid-19: Amazonas já transferiu 424 pacientes para outros estados – https://agenciabrasil.ebc.com.br/saude/noticia/2021-02/covid-19-amazonas-ja-transferiu-424-pacientes-para-outros-estados (2021).

41. S. L. Pond, S. D. Frost, S. V. Muse, HyPhy: hypothesis testing using phylogenies. Bioinformatics 21, 676–679 (2005).

42. X. X. Qu et al., Identification of two critical amino acid residues of the severe acute respiratory syndrome coronavirus spike protein for its variation in zoonotic tropism transition via a double substitution strategy. The Journal of Biological Chemistry 280, 29588–29595 (2005).

43. H. D. Song et al., Cross-host evolution of severe acute respiratory syndrome coronavirus in palm civet and human. Proceedings of the National Academy of Sciences of the United States of America 102, 2430–2435 (2005).

44. W. Li et al., Receptor and viral determinants of SARS-coronavirus adaptation to human ACE2. EMBO J 24, 1634–1643 (2005).

45. D. Zhou, Dejnirattisai, W., Supasa, P., Liu, C., Mentzer, A. J., Ginn, H. M., Zhao, Y., Duyvesteyn, H. M. E., Tuekprakhon, A., Nutalai, R., Wang, B., Paesen, G. C., Lopez-Camacho, C., Slon-Campos, J., Hallis, B., Coombes, N., Bewley, K., Charlton, S., Walter, T. S., Skelly, D., Lumley, S. F., Dold, C., Levin, R., Dong, T., Pollard, A. J., Knight, J. C., Crook, D., Lambe, T., Clutterbuck, E., Bibi, S., Flaxman, A., Bittaye, M., Belij-Rammerstorfer, S., Gilbert, S., James, W., Carroll, M. W., Klenerman, P., Barnes, E., Dunachie, S. J., Fry, E. E., Mongkolspaya, J., Ren, J., Stuart, D. I., Screaton, G. R., Evidence of escape of SARS-CoV-2 variant B.1.351 from natural and vaccine induced sera. Cell, (2021).

46. Z. Wang, Schmidt, F., Weisblum, Y., Muecksch, F., Barnes, C. O., Finkin, S., Schaefer-Babajew, D., Cipolla, M., Gaebler, C., Lieberman, J. A., Yang, Z., Abernathy, M. E., Huey-Tubman, K. E., Hurley, A., Turroja West, M. K. A., Gordon, K., Millard, K. G., Ramos, V., Da Silva, J., Xu, J., Colbert, R. A., Patel, R., Dizon, J., Unson-O’Brien, C., Shimeliovich, I., Gazumyan, A., Caskey, M., Bjorkman, P. J., Casellas, R., Hatziioannou, T., Bieniasz, P.D. Nussenzweig, M. C., mRNA vaccine-elicited antibodies to SARS-CoV-2 and circulating variants. Cold Spring Harbor Laboratory p. 2021.01.15.426911, (2021).

47. C. K. Wibmer, Ayres, F., Hermanus, T., Madzivhandila, M., Kgagudi, P., Lambson, B. E., Vermeulen, M., van den Berg, K., Rossouw, T., Boswell, M., Ueckermann, V., Meiring, S., von Gottberg, A., Cohen, C., Morris, L., Bhiman, J. N., Moore, P. L., SARS-CoV-2 501Y.V2 escapes neutralization by South African COVID-19 donor plasma. Cold Spring Harbor Laboratory p. 2021.01.18.427166, (2021).

48. S. Cele, Gazy, I., Jackson, L., Hwa, S.-H., Tegally, H., Lustig, G., Giandhari, J., Pillay, S., Wilkinson, E., Naidoo, Y., Karim, F., Ganga, Y., Khan, K., Balazs, A. B., Gosnell, B. I., Hanekom, W., Moosa, M. Y. S., NGS-SA, COMMIT-KZN Team, Lessells, R. J., Oliveira, T, Sigal, A., Escape of SARS-CoV-2 501Y.V2 variants from neutralization by convalescent plasma (available at https://www.krisp.org.za/publications.php?pubid=316). (2021).

49. L. Piccoli et al., Mapping Neutralizing and Immunodominant Sites on the SARS-CoV-2 Spike Receptor-Binding Domain by Structure-Guided High-Resolution Serology. Cell 183, 1024–1042 e1021 (2020).

50. E. Farfour et al., The Allplex 2019-nCoV (Seegene) assay: which performances are for SARS-CoV-2 infection diagnosis? European Journal of Clinical Microbiology & Infectious Diseases 39, 1997–2000 (2020).

51. F. M. Liotti et al., Evaluation of three commercial assays for SARS-CoV-2 molecular detection in upper respiratory tract samples. European Journal of Clinical Microbiology & Infectious Diseases 40, 269–277 (2021).

52. H. Li et al., The Sequence Alignment/Map format and SAMtools. Bioinformatics 25, 2078–2079 (2009).

53. I. Milne et al., Tablet--next generation sequence assembly visualization. Bioinformatics 26, 401–402 (2010).

54. R. C. Team. (R Foundation for Statistical Computing, Vienna, Austria, 2014).

55. M. A. Suchard et al., Bayesian phylogenetic and phylodynamic data integration using BEAST 1.10. Virus Evol 4, vey016 (2018).

56. D. L. Ayres et al., BEAGLE: an application programming interface and high-performance computing library for statistical phylogenetics. Systematic Biology 61, 170–173 (2012).

57. A. Rambaut, A. J. Drummond, D. Xie, G. Baele, M. A. Suchard, Posterior Summarization in Bayesian Phylogenetics Using Tracer 1.7. Systematic Biology 67, 901–904 (2018).

58. M. Worobey et al., 1970s and ‘Patient 0’ HIV-1 genomes illuminate early HIV/AIDS history in North America. Nature 539, 98–101 (2016).

59. M. Bletsa et al., Divergence dating using mixed effects clock modelling: An application to HIV-1. Virus Evol 5, vez036 (2019).

60. B. Murrell et al., Detecting individual sites subject to episodic diversifying selection. PLoS Genetics 8, e1002764 (2012).

61. O. A. MacLean et al., Evidence of significant natural selection in the evolution of SARS-CoV-2 in bats, not humans. bioRxiv (https://doi.org/10.1101/2020.05.28.122366, 2020).

62. A. G. Wrobel et al., SARS-CoV-2 and bat RaTG13 spike glycoprotein structures inform on virus evolution and furin-cleavage effects. Nat Struct Mol Biol 27, 763–767 (2020).

63. L. Schrödinger. The PyMOL Molecular Graphics, version 2.0 (2015).

64. S. Flaxman et al., Estimating the effects of non-pharmaceutical interventions on COVID-19 in Europe. Nature 584, 257–261 (2020).

65. R. Bellman, Harris, T., On Age-Dependent Binary Branching Processes. In: The Annals of Mathematics (1952).

66. W. Feller, On the Integral Equation of Renewal Theory. (In: The Annals of Mathematical Statistics, 1941).

67. N. Brazeau, Verity, R., Jenks, S., Fu, H., Whittaker, C., Winskilll, P., Dorigatti, I., Walker, P., Riley, S., Schenekenberg, R. P., et al., “Report 34: COVID-19 infection fatality ratio: estimates from seroprevalence” (https://www.imperial.ac.uk/mrc-global-infectious-disease-analysis/covid-19/report-34-ifr/, 2020).

68. E. Volz, Mishra, S., Chand, M., Barrett, J. C., Johnson, R., Geidelberg, L., Hinsley, W. R., Laydon, D. J., Dabrrera, G., O’Toole, A., Amato, R., Ragonnet-Crronin, M., Harrison, I., Jackson, B., Ariaani, C. V., Boyd, O., Loman, N. J., McCrone, J. T., Goncalves, S., Jorgensen, D., Myers, R., Hill, V., Jackson, D. K., Gaythorpe, K., Groves, N., Sillitoe, J., Kwiatkowski, D. P., The COVID-19 Genomics UK (COG-UK) consortium, Flaxman, S., Ratmann, O., Bhatt, S., Hopkins, S., Gandy, A., Rambaut, A., Ferguson, N. M., Transmission of SARS-CoV-2 Lineage B.1.1.7 in England: Insights from linking epidemiological and genetic data. medRxiv (https://doi.org/10.1101/2020.12.30.20249034, 2021).

69. T. Mellan, Hoeltgebaum, H., Mishra, S., Whittaker, C., Schnekenberg, R. P,, A. Gandy, Unwin, H. J. H., Vollmer, M. A., Coupland, H., Hawryluk, I., Faria, N. R., Vesga, J., Zhu, H., Hutchinson, M., Ratmann, O., Monod, M.,, K. Ainslie, Baguelin, M., Bhatia, S., Boonyasiri, A., Brazeau, N., Charles, G., Cooper, L. V., Cucunuba, Z., Cuomo-Dannenburg, G., Dighe, A., Djaafara, B., Eaton, J., van Elsland, S. L., FitzJohn, R., Fraser, K., Gaythorpe, K., Green, W., Hayes, S., Imai, N., Jeffrey, B., Knock, E., Laydon, D., Lees, J., Mangal, T., Mousa, A., Nedjati-Gilani, G.,, P. Nouvellet, Olivera, D., Parag, K. V., Pickles, M., Thompson, H. A., Verity, R., Walters, C., Wang, H., Wang, Y., Watson, O. J., Whittles, L., Xi, X., Okell, L., Dorigatti, I., Walker, P., Ghani, A., Riley, S., Ferguson, N., Donnelly, C. A., Flaxman, S., Bhatt, S., Report 21: Estimating COVID-19 cases and reproduction number in Brazil. Imperial College London (08-05-2020)*, doi:* https://doi.org/10.25561/78872., (2020).

70. I. Hawryluk et al., Inference of COVID-19 epidemiological distributions from Brazilian hospital data. Journal of the Royal Society 17, 20200596 (2020).

71. DATASUS, Ministry of Health, SRAG 2020—severe acute respiratory syndrome database—including data from COVID-19. Surveillance of severe acute respiratory syndrome (SARS) https://opendatasus.saude.gov.br/dataset/bd-srag-20210 (2020).

72. DATASUS, Ministry of Health, SRAG 2021—severe acute respiratory syndrome database—including data from COVID-19. Surveillance of severe acute respiratory syndrome (SARS) https://opendatasus.saude.gov.br/dataset/bd-srag-2021 (2021).

73. J. Hellewell, Russell, T. W., Beale, R., Kelly, G., Houlihan, C., Nastouli, E., Kucharski, J., SAFER Investigators, Field Study Team, Crick COVID-19 Consortium, et al., Estimating the effectiveness of routine asymptomatic PCR testing at different frequencies for the detection of SARS-CoV-2 infections. medRxiv (https://doi.org/10.1101/2020.11.24.20229948, 2020).

74. B. Borremans, Gamble, A., Prager, K. C., Helmaan, S. K., McClain, A. M., Cox, C., Savage, V., O’Lloyd-Smith, J., Quantifying antibody kinetics and RNA detection during early-phase SARS-CoV-2 infection by time since symptom onset. eLife e60122, (2020).

75. Fundação de Vigilância em Saúde do Amazonas COVID-19 no Amazonas, “Dados epidemiológicos e financeiros das ações de combate à COVID-19. Publicações”

76. Y. Shu, J. McCauley, GISAID: Global initiative on sharing all influenza data - from vision to reality. Euro Surveillance 22, (2017).

